# Determinants of Obesity in West Africa: A Systematic Review

**DOI:** 10.1101/2021.04.27.21255462

**Authors:** Kingsley Agyemang, Subhash Pokhrel, Christina Victor, Nana Kwame Anokye

**Author notes:** Corresponding author Kingsley Agyemang Division of Global Public Health Department of Health Sciences College of Health, Medicine, and Life Sciences Brunel University London.

## Abstract

**Objectives:** Obesity prevalence is increasing in West Africa. This study explores obesity determinants in West Africa to inform policy.

**Methods:** Scopus, Web of Science and PsycINFO were searched for relevant papers from March to April 2020. The search strategy included combinations of key words specific to each database. Eligibility criteria included studies on obesity determinants conducted in West Africa, and involving participants aged eighteen years and above. The quality of the studies was appraised using the Agency for Healthcare Research and Quality checklist. Data was synthesized qualitatively.

**Results:** Sixty-three (63) papers were selected. Majority of the studies originated from Ghana (n=22) and Nigeria (n=19). All included studies used cross-sectional study design. In all, 36 determinants were identified, of which 20 were demographic, socio-economic, lifestyle and biological factors, and sixteen 16 were environmental factors, like physical proximity to fast food outlets. Increasing age (OR=0.09, 95% CI= 0.12 to 65.91) and being a woman (OR=1.38, 95% CI=1.18 to 55.40) were the common determinants of obesity in West Africa.

**Conclusion:** Obesity in West Africa is determined by complex multi-faceted factors. There is an urgent need for robust engagement with wider stakeholder groups to develop obesity prevention and control policies in West Africa.

## Introduction

Obesity is a global public health concern, and the World Health Organization (WHO) has estimated that it affects 650 million adults worldwide, with this burden projected to increase to one billion globally by 2030^1, 2^. It is a modifiable risk factor for non-communicable diseases (NCDs), such as cardiovascular disease, type 2 diabetes, and various cancers^2, 3^. In 2018, NCDs were responsible for 41 million (71%) of the world’s 58 million deaths, with 15 million dying prematurely^4^. Low- and middle-income countries, including Africa, bear over 85% of the burden of these NCDs premature deaths, resulting in cumulative economic losses of US$7 trillion over the next 15 years and millions of people trapped in poverty^5^. Specifically, deaths from NCDs are likely to increase by 27% (about 28 million additional deaths) in Africa and are projected to exceed other deaths combined by 2030 in the region countries^6^.

Until recently, Africa was minimally affected by the obesity epidemic due to the burden of under-nutrition, HIV/AIDs, and tuberculosis^7^. However, current evidence shows that the continent is experiencing a swift rise in overweight and obesity prevalence as well as its associated co-morbidities because of a rapid ongoing nutritional and epidemiological transitions^7, 8, 9^. These dynamics, coupled with the epidemic of NCDs, create an urgent need for evidence-based sustainable actions to mitigate the obesity burden in Africa.

In West Africa, one of WHO’s Africa subregions, about 52million people are with obesity, making it one of the subregions burdened with the obesity epidemic^10^. Based on this, obesity prevention and control policies have been implemented to address the epidemic. Key among them is the *National Policy for the Prevention and Control of Chronic Non-Communicable Diseases (NCDs) in Ghana, the National Policy on Food and Nutrition (NPFN) in Nigeria and the National Food and Nutrition Security (NFNS) in Sierra Leone*. Although these policies have created awareness about the obesity problem in West Africa, the prevalence of obesity continues to increase exponentially^11, 12^. One primary reason for the shortfall of the obesity policies in West Africa is its inability to capture, holistically, all the determining factors of obesity^13, 14^. Also, there is limited evidence to guide the implementation of these policies.

Several obesity determinants that could inform obesity prevention and control policies are identified in the literature. However, most of these determinants were discovered in high-income-countries (HICs); thus, they do not provide setting-specific obesity interventions/policies for West African countries. Hence, there is an unmet demand for a review of the determinants of obesity in West Africa. Therefore, this study aims to review factors associated with obesity in West Africa to inform context-specific interventions to address the obesity epidemic on the subregion. The findings may also provide recommendations for future empirical research in other LMICs.

## Methods

### Search strategy

Three electronic databases, Scopus, Web of Science and PsycINFO, were searched from March to April 2020 (updated search conducted from January to February 2021) for eligible studies using all possible combinations of search terms related to obesity determinants and Africa (Table 1). References of the papers identified in the electronic search were also screened for relevant studies. Identified studies were screened against the following eligibility criteria: (a)Quantitative studies that explored the determinants of obesity, (b) Studies written in the English language given the limited resources for translation, (c) Studies with participants aged eighteen years and above, and (d) Studies that used samples from West Africa settings.

**Table 1:**
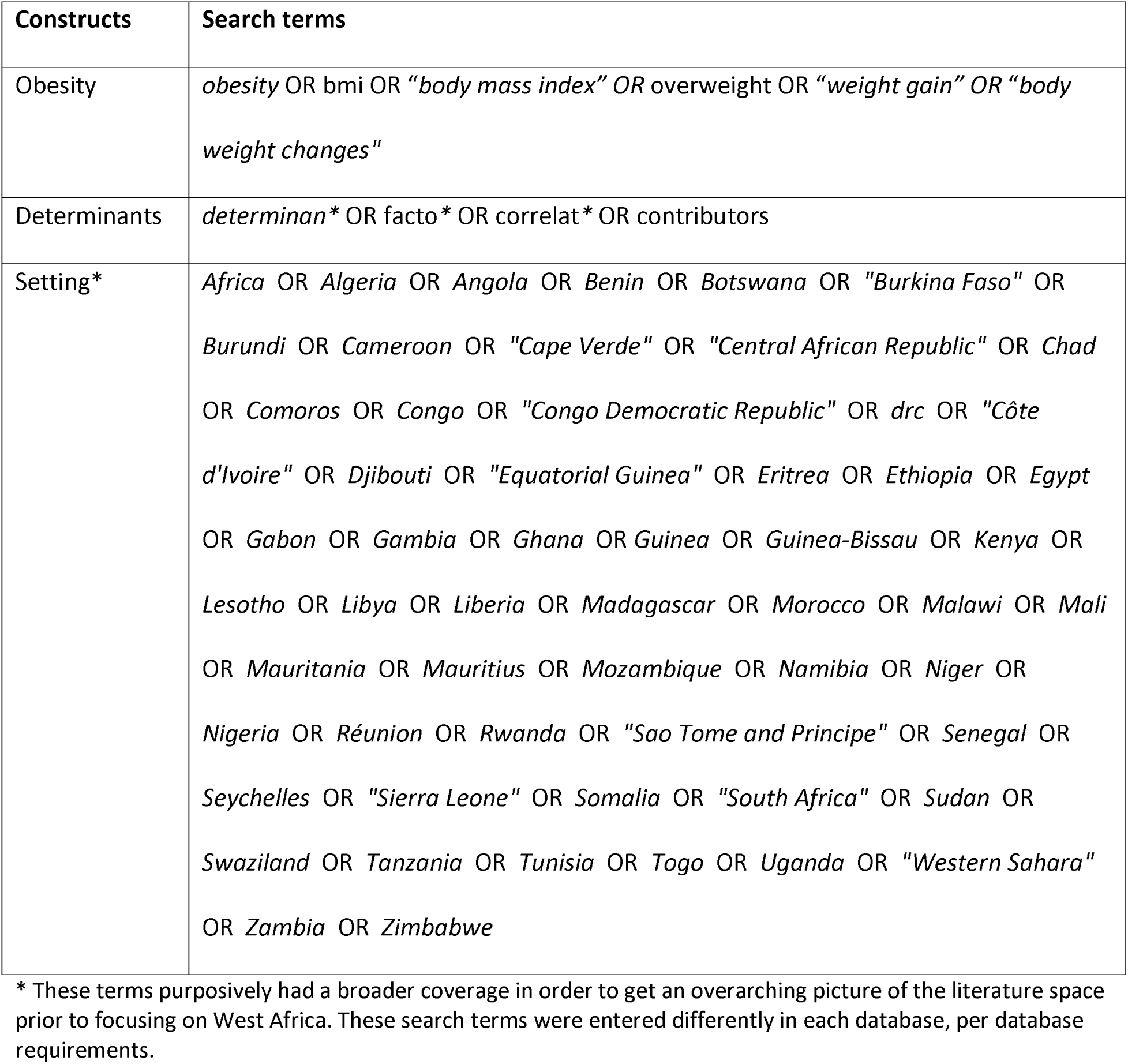
Search terms

| Constructs | Search terms |
| --- | --- |
| Obesity | *obesity OR bmi OR "body mass index" OR overweight OR "weight gain" OR "body weight changes"* |
| Determinants | *determinan\* OR facto\* OR correlat\* OR contributors* |
| Setting* | *Africa OR Algeria OR Angola OR Benin OR Botswana OR "Burkina Faso" OR Burundi OR Cameroon OR "Cape Verde" OR "Central African Republic" OR Chad OR Comoros OR Congo OR "Congo Democratic Republic" OR drc OR "Côte d'Ivoire" OR Djibouti OR "Equatorial Guinea" OR Eritrea OR Ethiopia OR Egypt OR Gabon OR Gambia OR Ghana OR Guinea OR Guinea-Bissau OR Kenya OR Lesotho OR Libya OR Liberia OR Madagascar OR Morocco OR Malawi OR Mali OR Mauritania OR Mauritius OR Mozambique OR Namibia OR Niger OR Nigeria OR Réunion OR Rwanda OR "Sao Tome and Principe" OR Senegal OR Seychelles OR "Sierra Leone" OR Somalia OR "South Africa" OR Sudan OR Swaziland OR Tanzania OR Tunisia OR Togo OR Uganda OR "Western Sahara" OR Zambia OR Zimbabwe* |

\* These terms purposively had a broader coverage in order to get an overarching picture of the literature space prior to focusing on West Africa. These search terms were entered differently in each database, per database requirements.

### Data extraction

Two independent reviewers extracted relevant data from the selected studies using standardized data extraction questions developed a priori. The data extraction questions included information on the general characteristics of the studies, such as authors name and year of publication; methodological features like sample size and sample characteristics; and findings and recommendations from the authors. The questions were reviewed and agreed on by all the authors and pilot-tested before their usage. To ensure the quality of data extraction, 50% of the included papers were randomly selected and reviewed by a third reviewer. Any disagreements above 5% were resolved by consensus or through the opinion of another independent reviewer.

### Quality appraisal

Two reviewers appraised the quality of the selected papers using the Agency for Healthcare Research and Quality (AHRQ) methodology Checklist to reflect the cross-sectional design of all included studies. The AHRQ assessed the quality of the included studies based on eleven checklists. For each question, a score of one (1) was given if the study met it and zero (0) if the study did not meet it. The scores of the questions were summed to represent the total score for each study. The total score for meeting all the criteria on the AHRQ checklist was eleven. None of the studies was classified as high quality since none of them met all the eleven checklists. Their scores ranged from 4 to 9, indicating a low to medium quality. Table 2 shows the number of studies that met each of the AHRQ checklist criteria.

**Table 2:** Number of studies that met the AHRQ checklists

| **AHRQ Checklists** | **Number/proportion (%) of studies that met it** |
| --- | --- |
| **1) Define the source of information (survey, record review)** | 61 (97) |
| **2) List inclusion and exclusion criteria for exposed and unexposed subjects (cases and controls) or refer to previous publications** | 50 (79) |
| **3) Indicate time period used for identifying patients** | 47 (75) |
| **4) Indicate whether or not subjects were consecutive if not population-based** | 44 (70) |
| **5) Indicate if evaluators of subjective components of study were masked to other aspects of the status of the participants** | 23 (37) |
| **6) Describe any assessments undertaken for quality assurance purposes (e.g., test/retest of primary outcome measurements)** | 9 (14) |
| **7) Explain any patient exclusions from analysis** | 17 (27) |
| **8) Describe how confounding was assessed and/or controlled.** | 22 (35) |
| **9) If applicable, explain how missing data were handled in the analysis** | 12 (19) |
| **10) Summarize patient response rates and completeness of data collection** | 31 (49) |
| **11) Clarify what follow-up, if any, was expected and the percentage of patients for which incomplete data or** | 8 (13) |

| AHRQ Checklists | Number/proportion (%) of studies that met it |
| --- | --- |
| follow-up was obtained |  |

### Data Synthesis/Analysis

Descriptive synthesis of data was performed to describe the methods, operationalization of methods, major limitations, suggestions, and recommendations for future research. Where appropriate, for each of these domains, the synthesis compared the findings from across the studies so that the themes could be put into wider perspectives.

## Results

A total of 4,085 records were identified from the database searches; however, only sixty-three met the inclusion criteria (figure 1). All the sixty-three papers were cross-sectional studies. Majority of them used primary data (n=44) while the remaining used secondary data. Structured questionnaires were the common instruments used in collecting the data. The sample size of the studies that used primary data ranged from 59 to 6,959, and that of studies with secondary data ranged from 600 to 1,225,816. Fifty-five of the studies focused on only one West African country, mainly Ghana (n=22), Nigeria (n=19), Cameroon (n=4), Burkina Faso (n=3), Cote d’ Ivoire (n= 2), Senegal (n=2), Togo (n=1), Gambia (n=1), Benin (n=1) while eight of them focused on a combination of West African countries like Mali, Sierra Leone, Guinea, Liberia and Niger. All the studies were published between 2003 and 2019, with most of them in 2016 (n=10) and 2017 (n=10).

**Figure 1:**
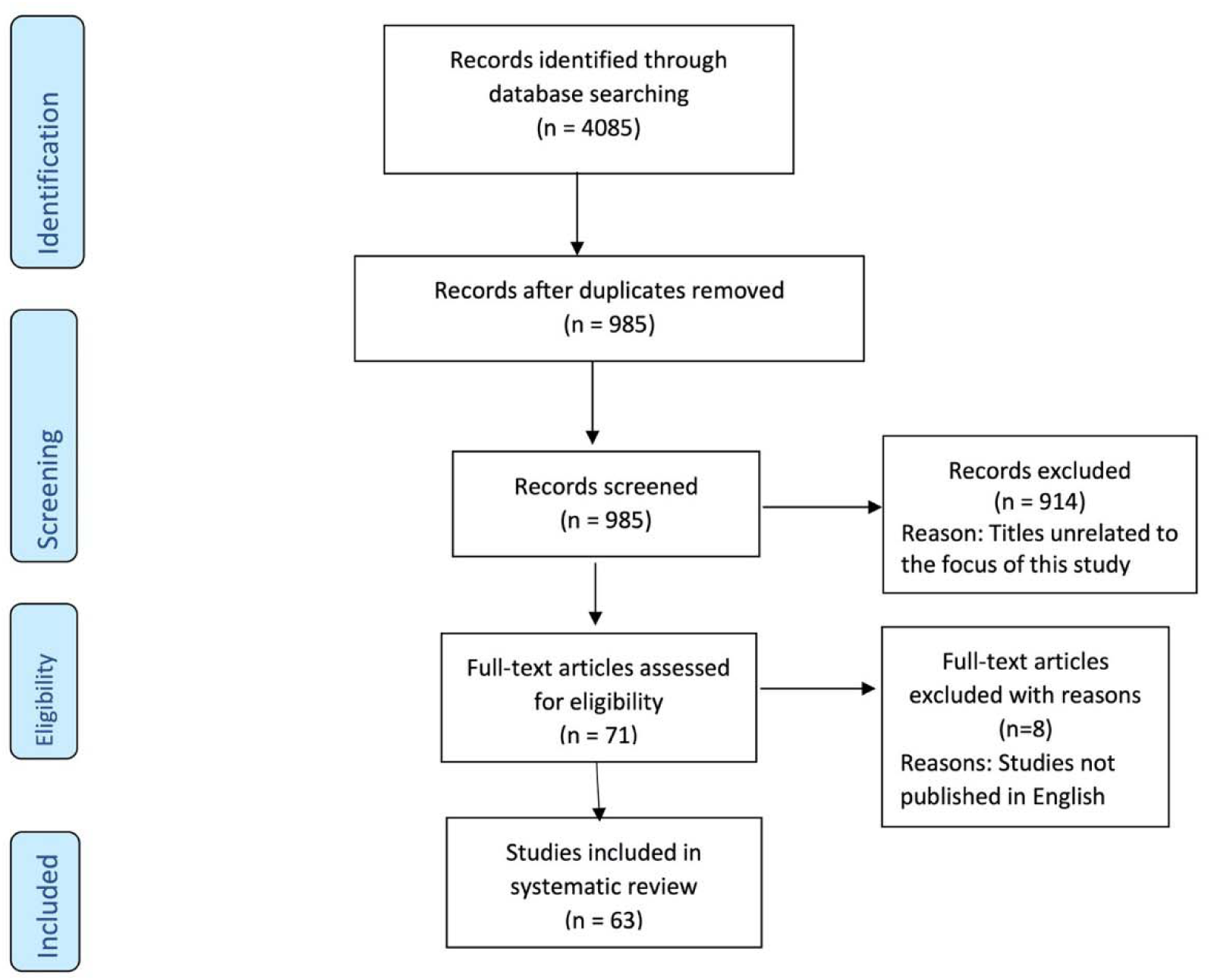
Prisma Diagramfor the Selection Process

### Measurement of determining variables

This review identified several determinants in the included studies. These determinants are categorised into demographic, socio-economic, biological, lifestyle and environmental variables, based on the Dahlgren and Whitehead ecological model of public^15^. The demographic variables included age and sex; socio-economic variables included level of education and employment status; biological variables comprised presence of hypertension, diabetes and hypercholesterolemia; lifestyle variables encompassed cigarette smoking, physical activity score, dietary score and alcohol consumption; and the environmental variables focused on neighborhood characteristics, like access to convenient stores and presence of recreational facilities, and extent of globalisation-which was an aggregate of social, political and economic globalisation that the study participants were exposed to.

### Definition/Measurement of obesity

All sixty-three studies defined obesity based on the WHO BMI measurement of obesity. Thus, they classified BMI of < 18.5 kg/m^2^ as people with underweight, 18.5–25 kg/m^2^ as people with normal weight, 25.0–29.9 kg/m^2^ as people with overweight and BMI ≥ 30.0 kg/m^2^ as people with obesity. Additionally, three of the studies further measured obesity body fat^16^ and abdominal obesity^17, 18^. Percentage body fat (BF %) was estimated using the following formula Adult body fat% = (1.20 × BMI) + (0.23 × Age) -(10.8 × sex) -5.4. The cut-offs for BF% for men and women were 25% and 30%, respectively. Abdominal obesity was classified as a waist circumference of ≥ 102 cm in men or ≥88 cm in women, per WHO definition. Among the 44 papers that collected primary data, 32 of them used self-reported height and weight measures to estimate the BMI of the participants. Table 3 and 4 provide summary of the study characteristics, including measured variables.

**Table 3:**
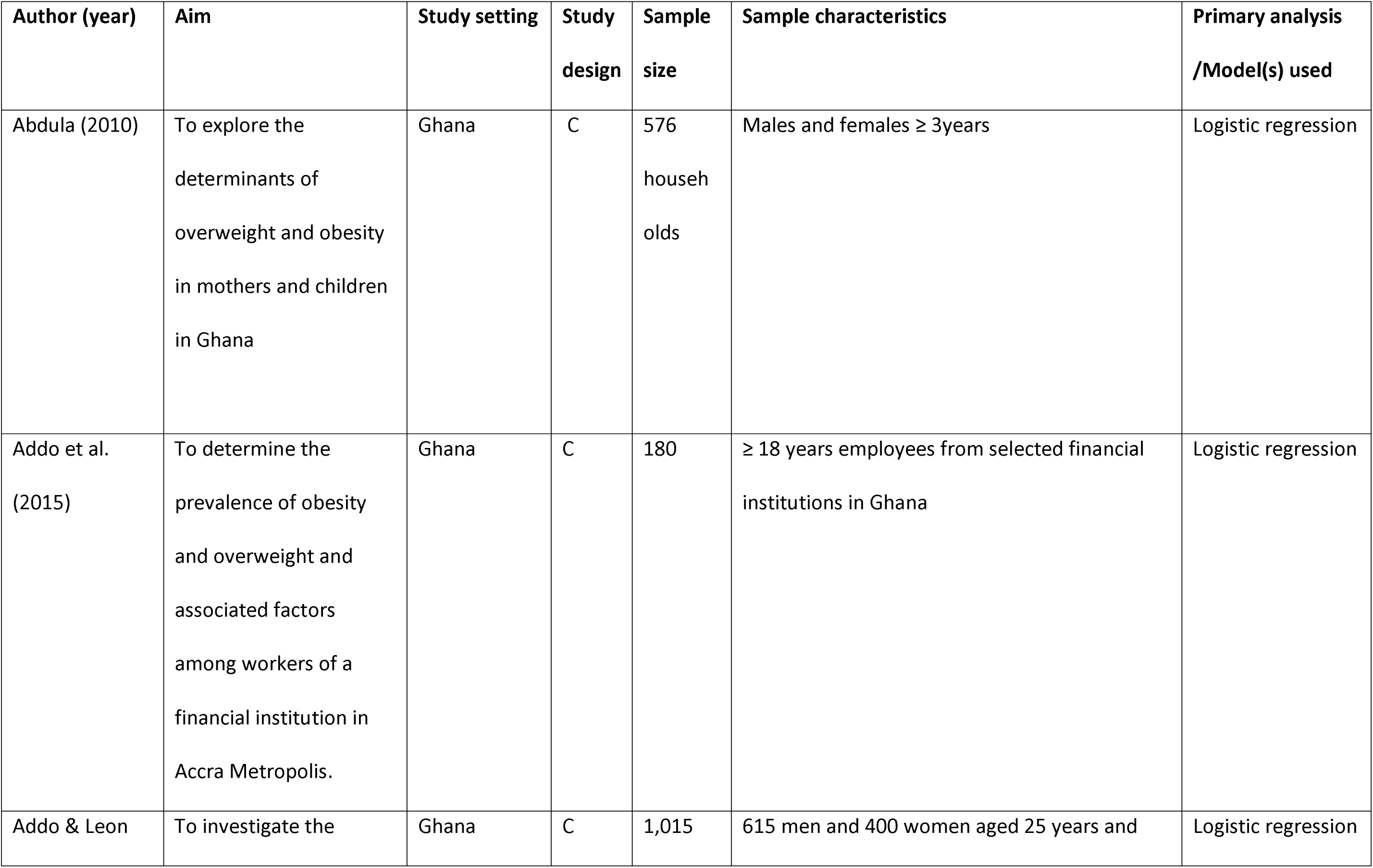
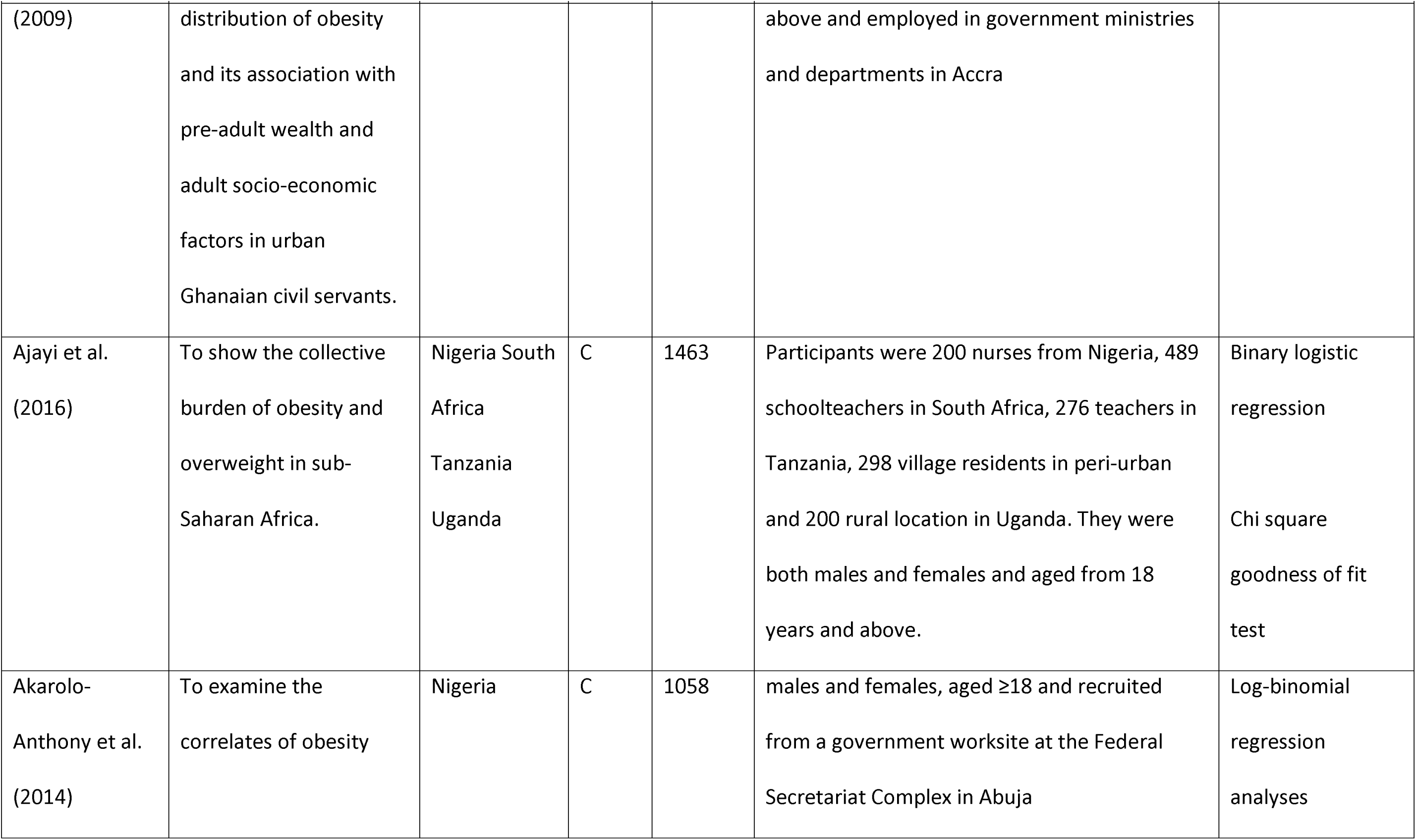
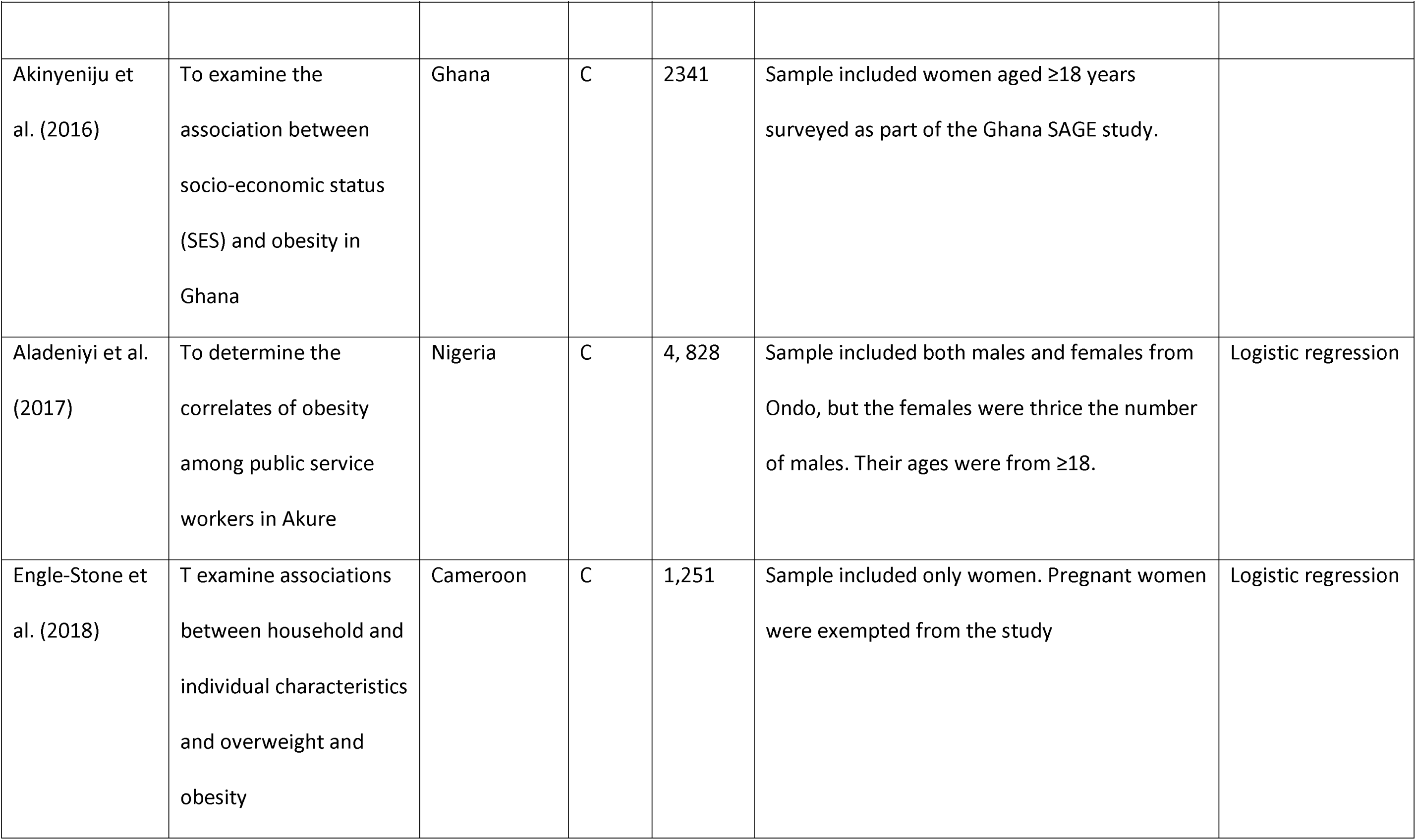
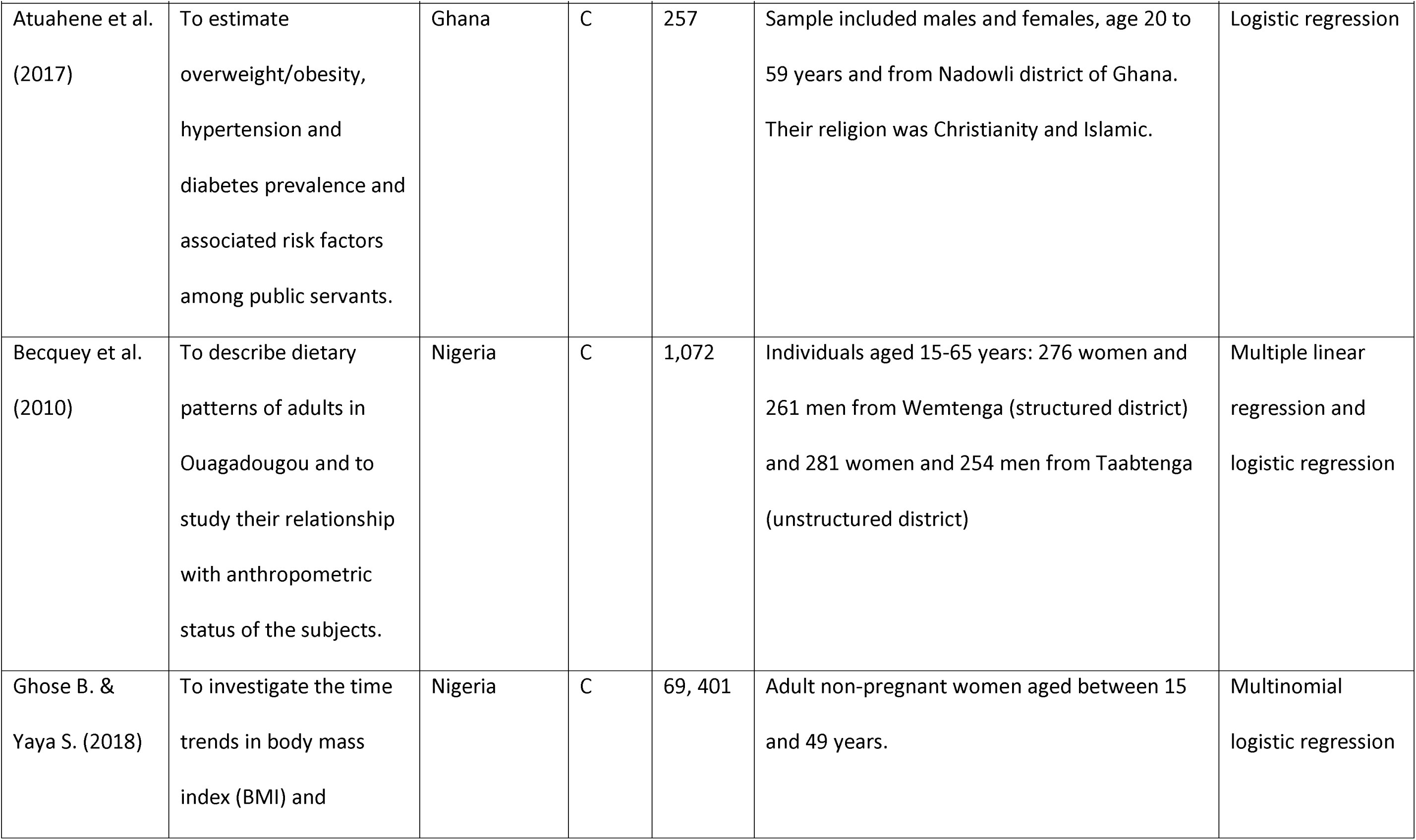
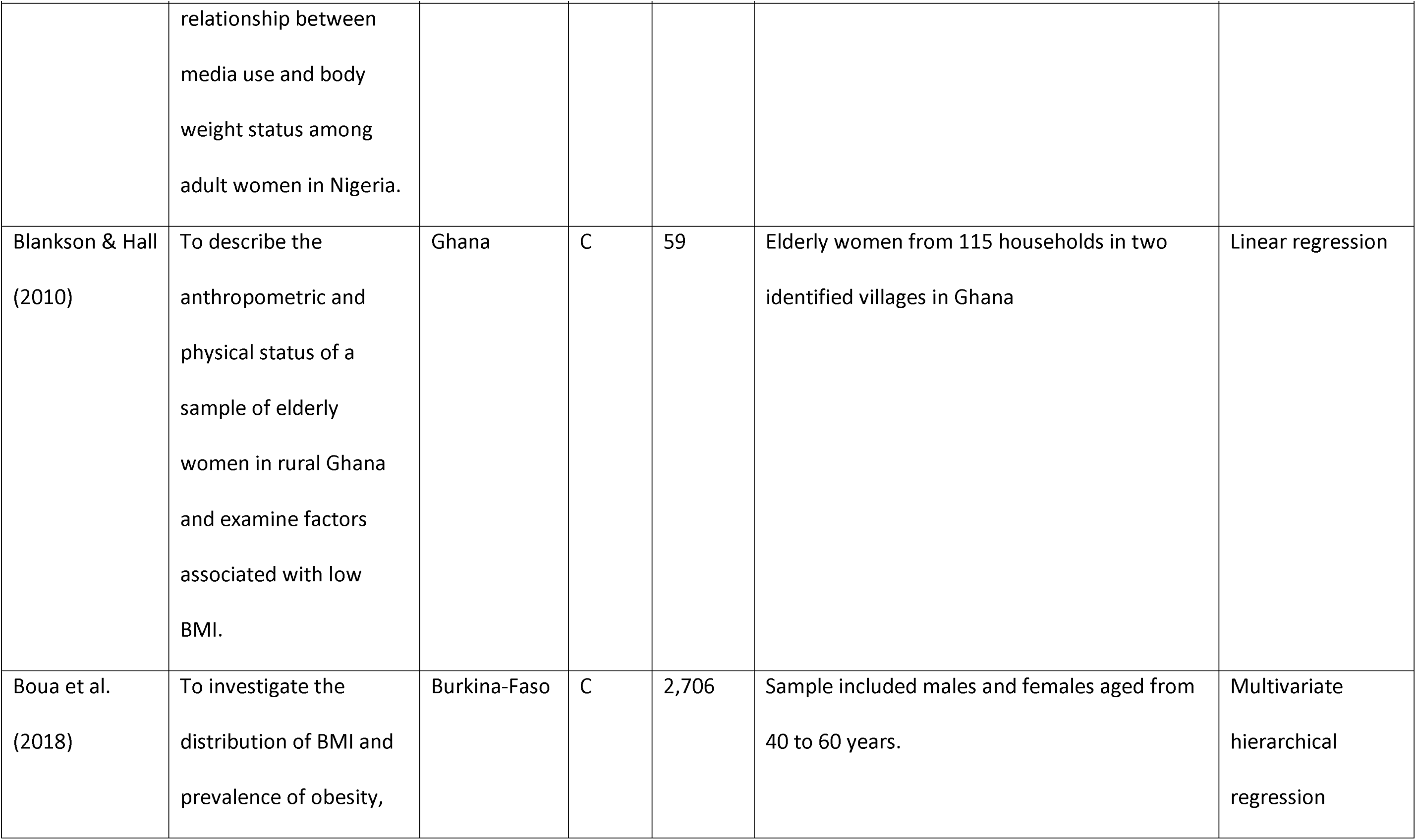
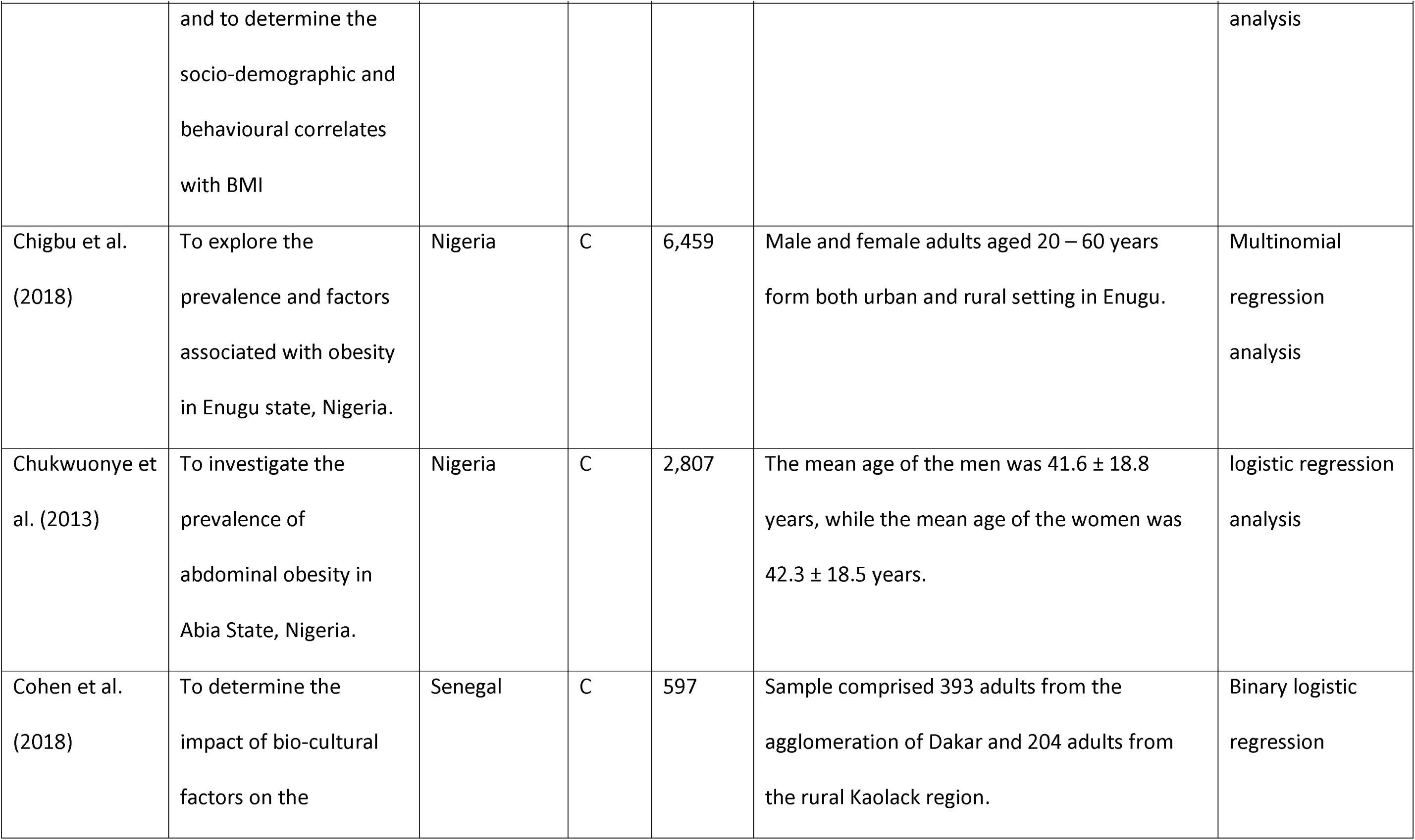
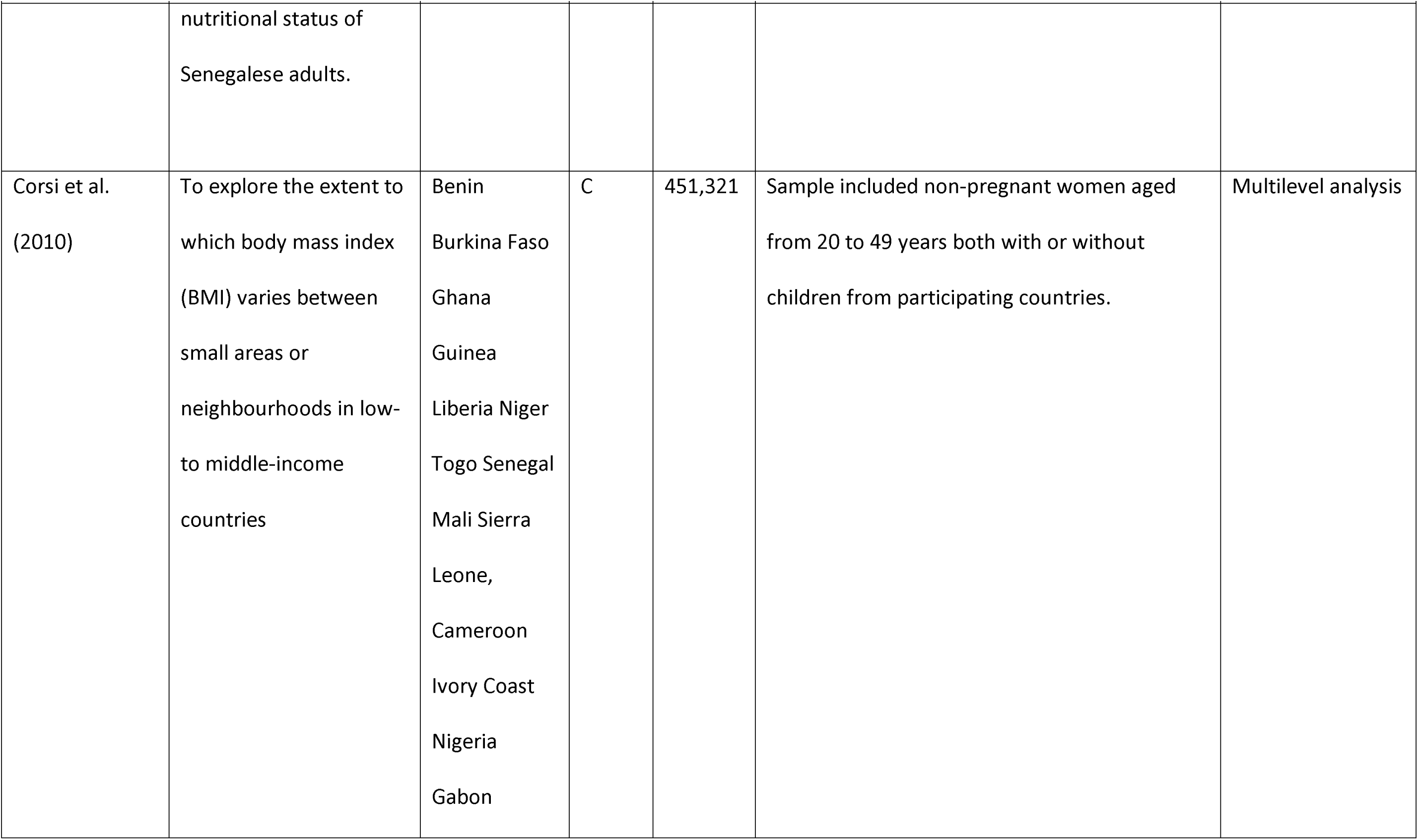
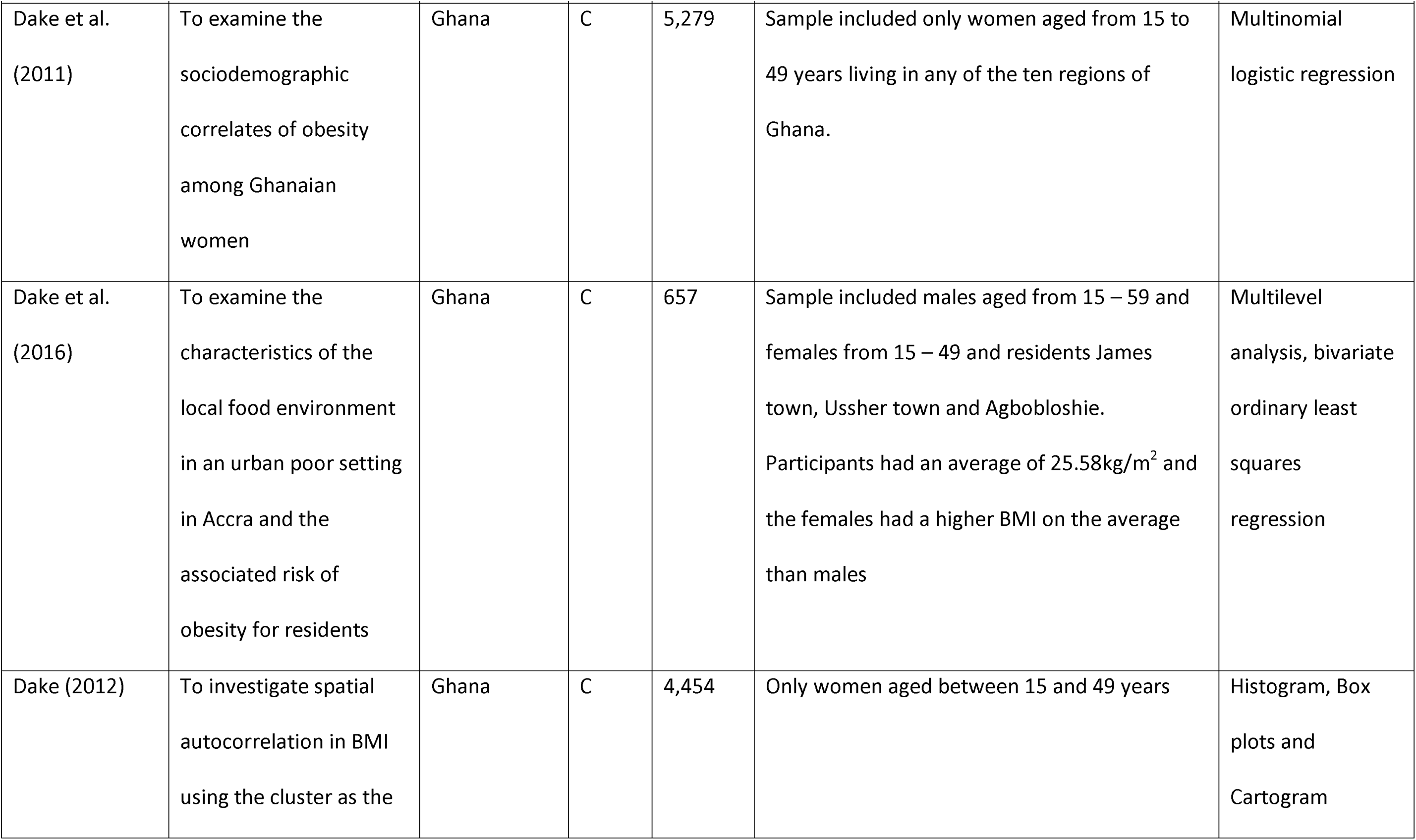
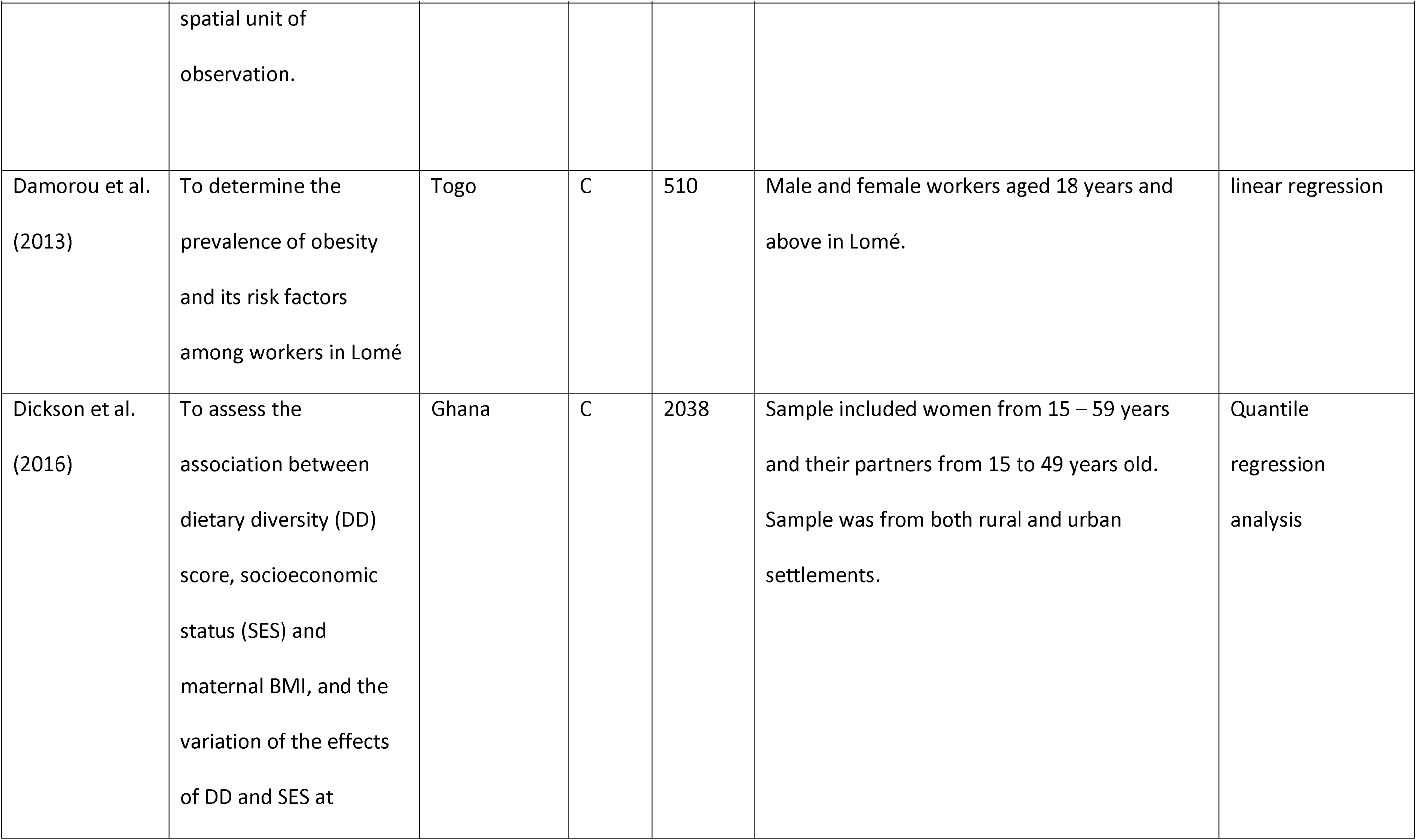
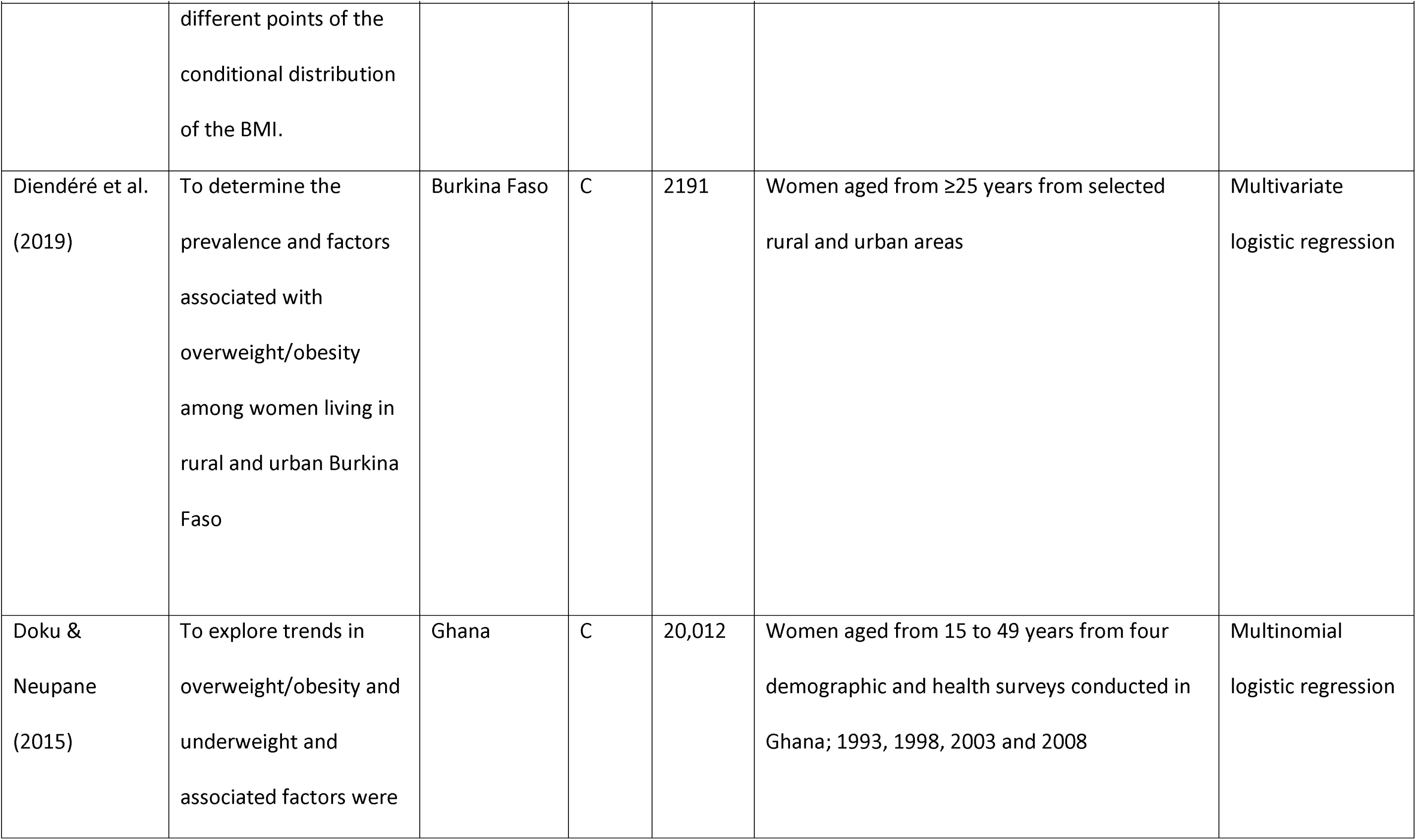
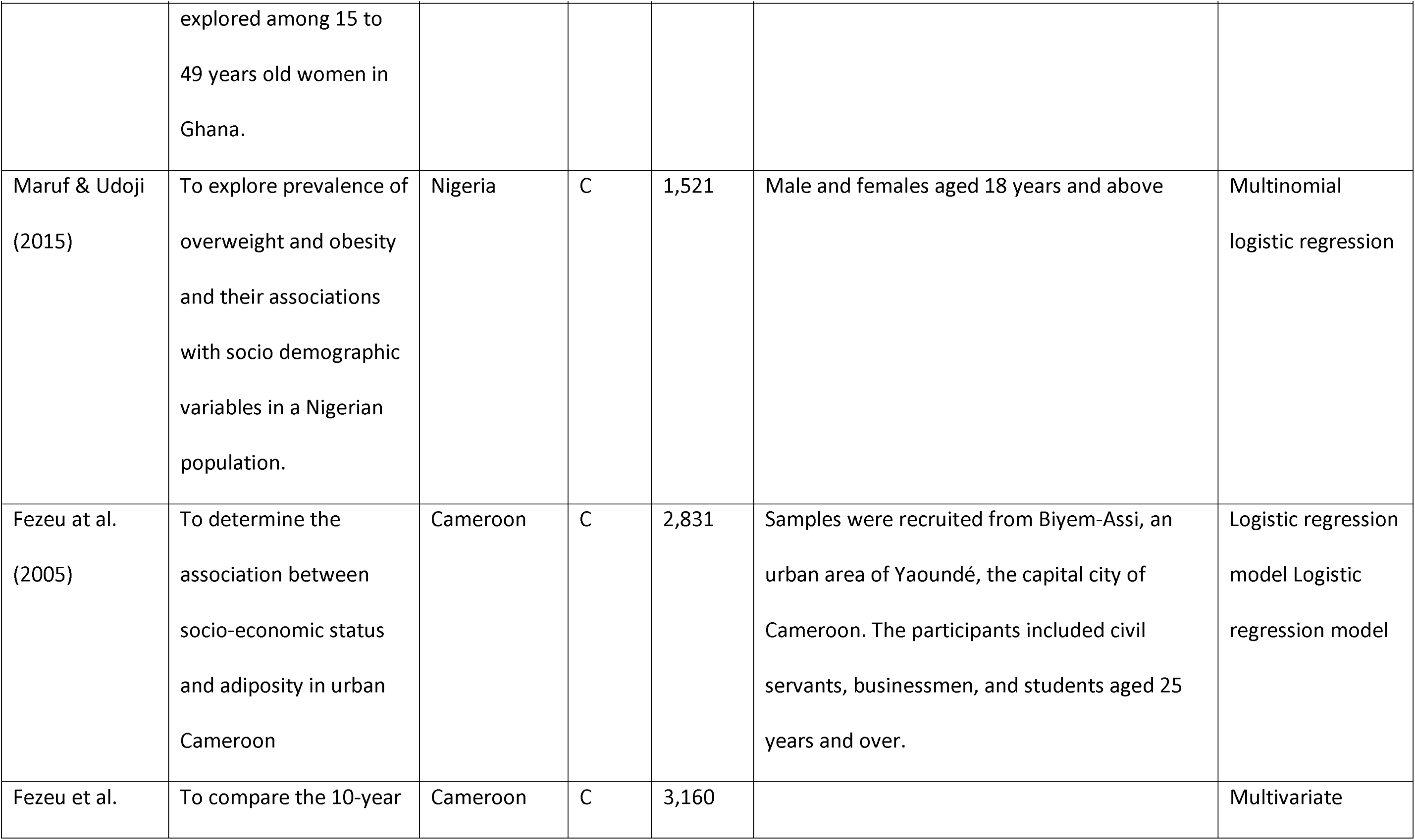
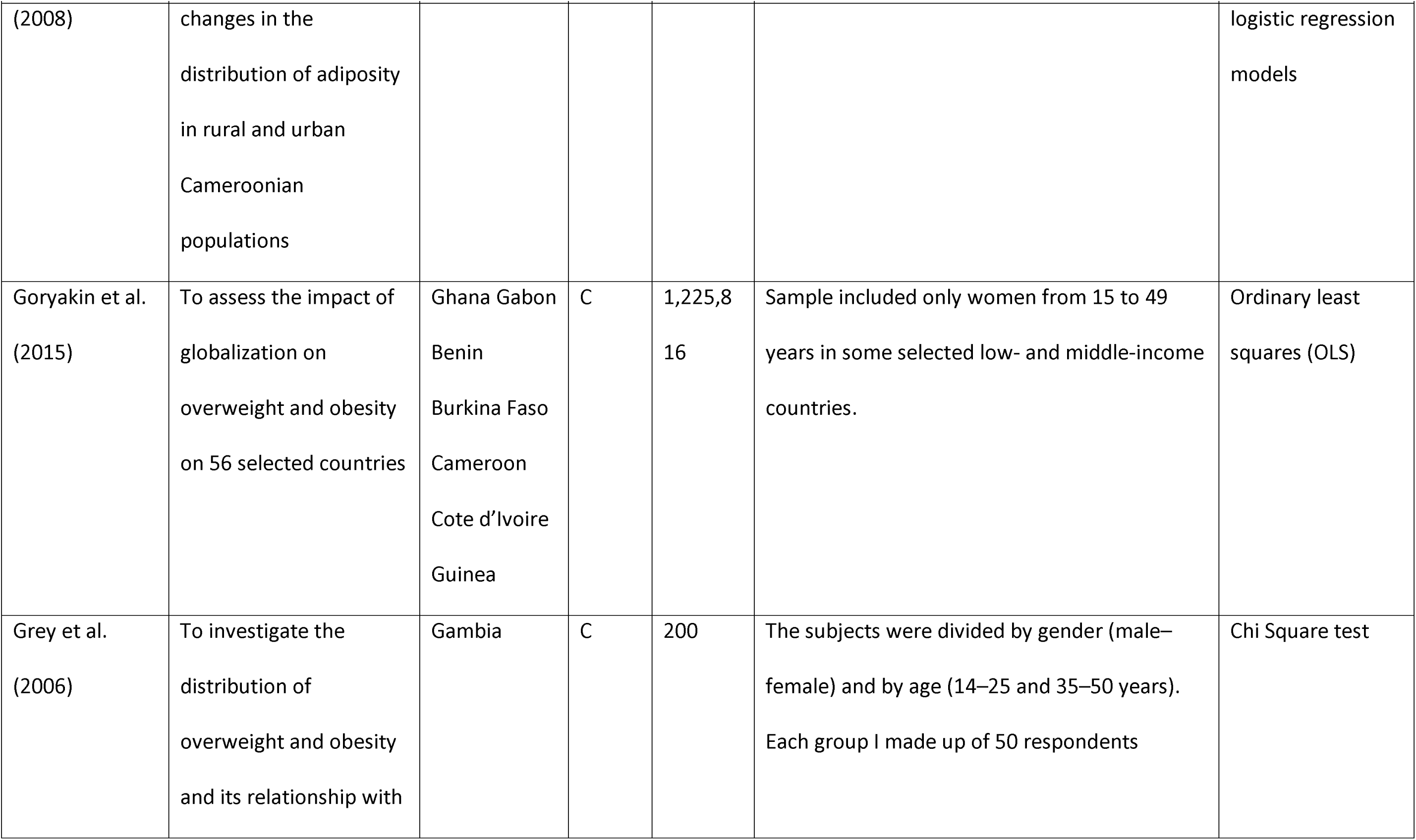
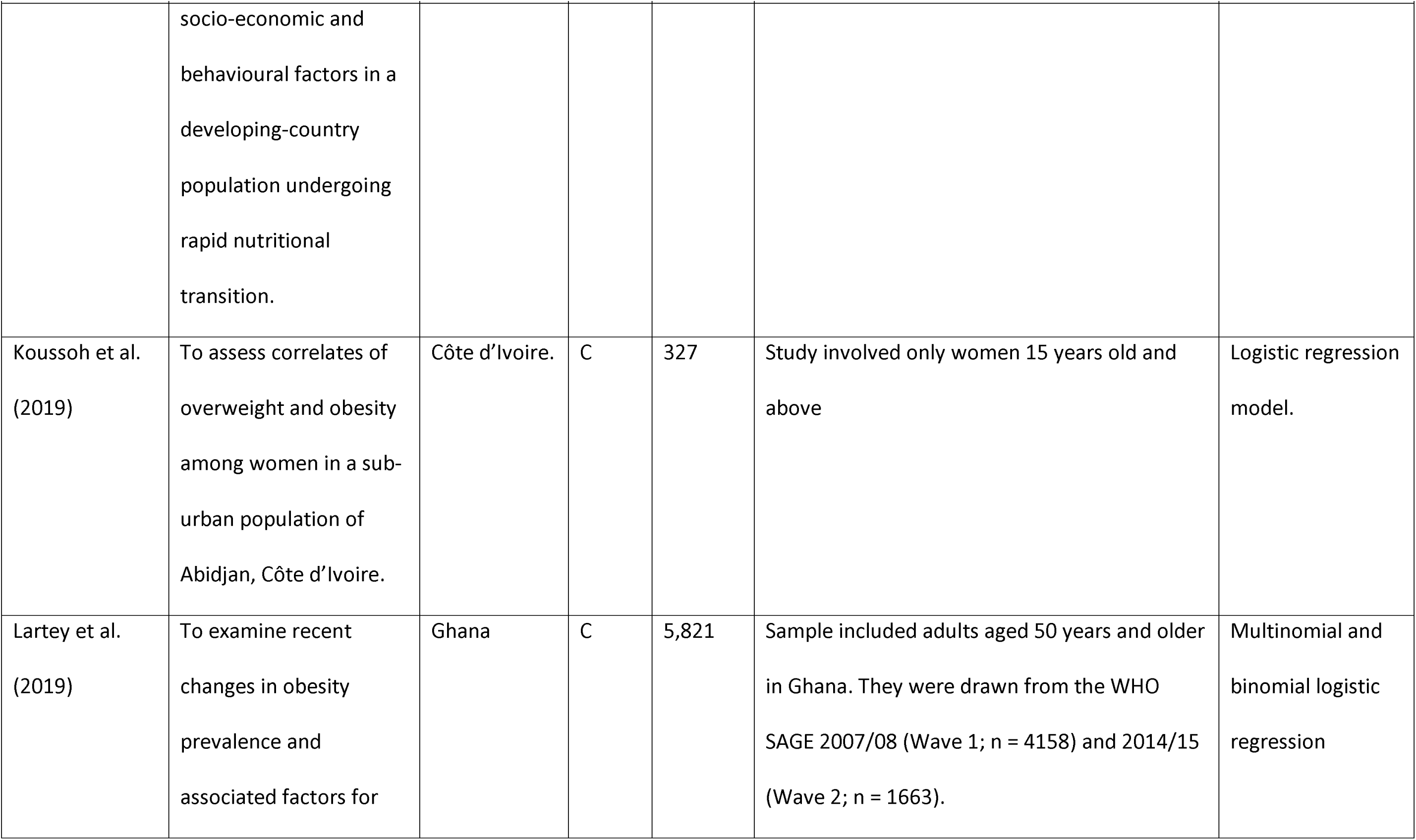
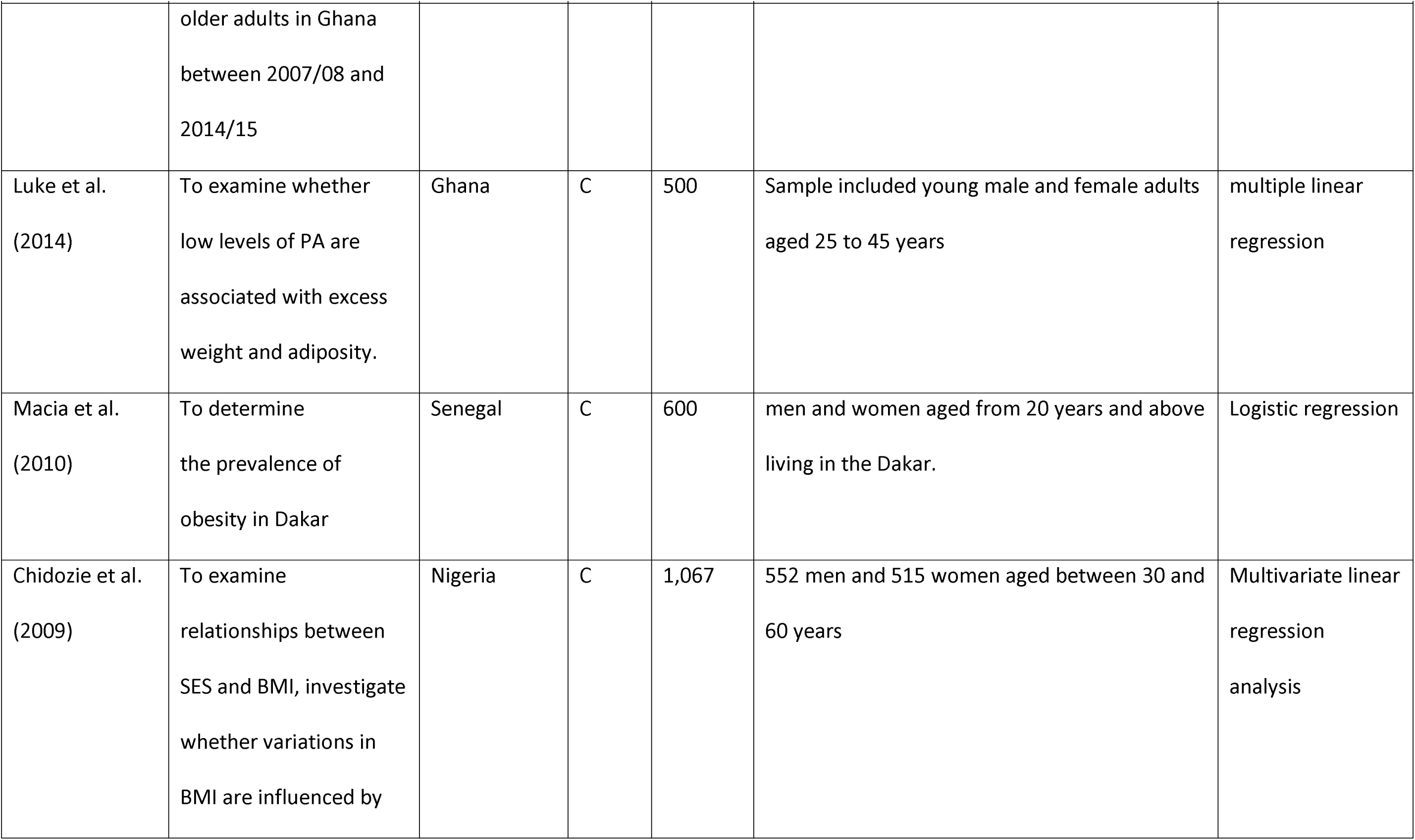
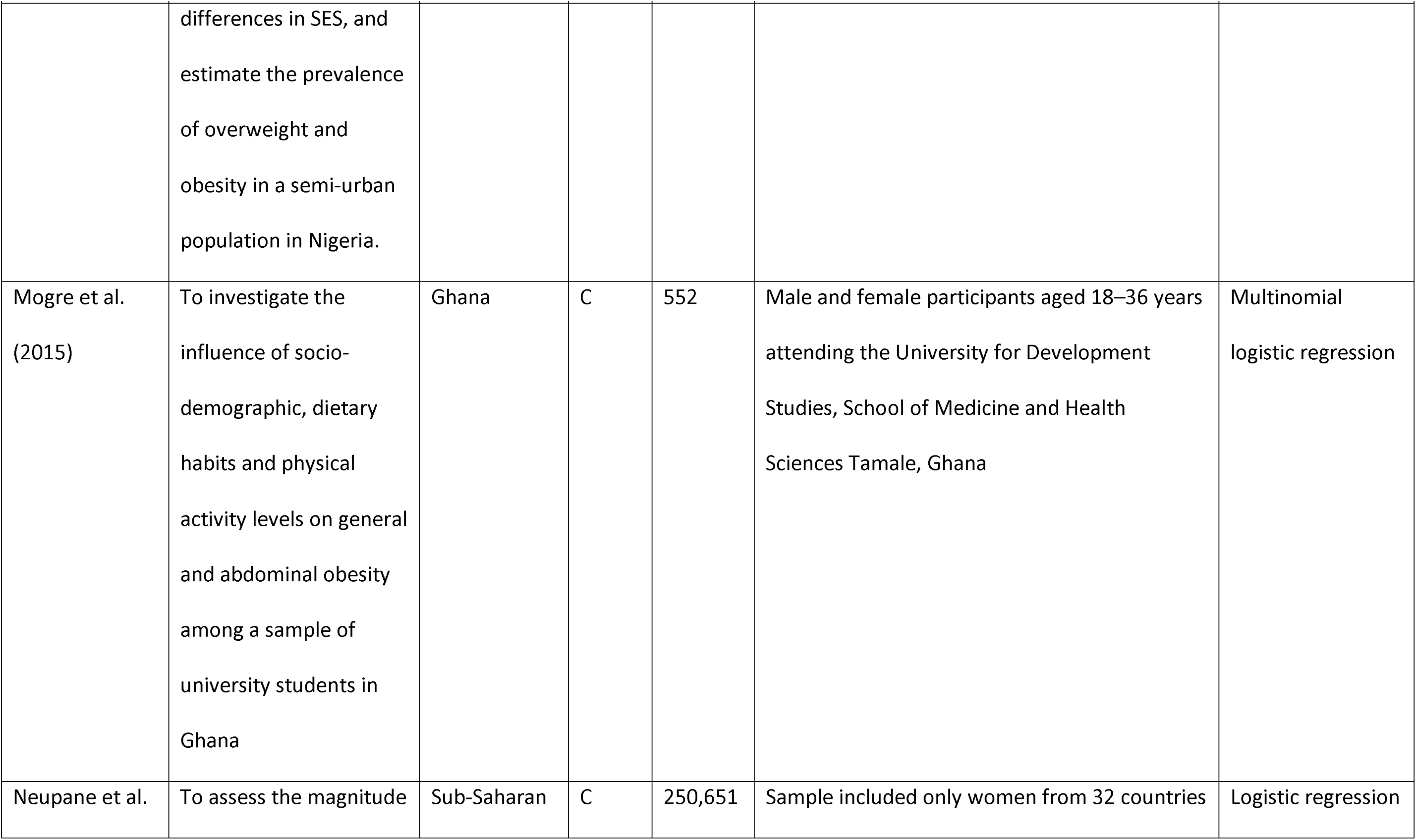
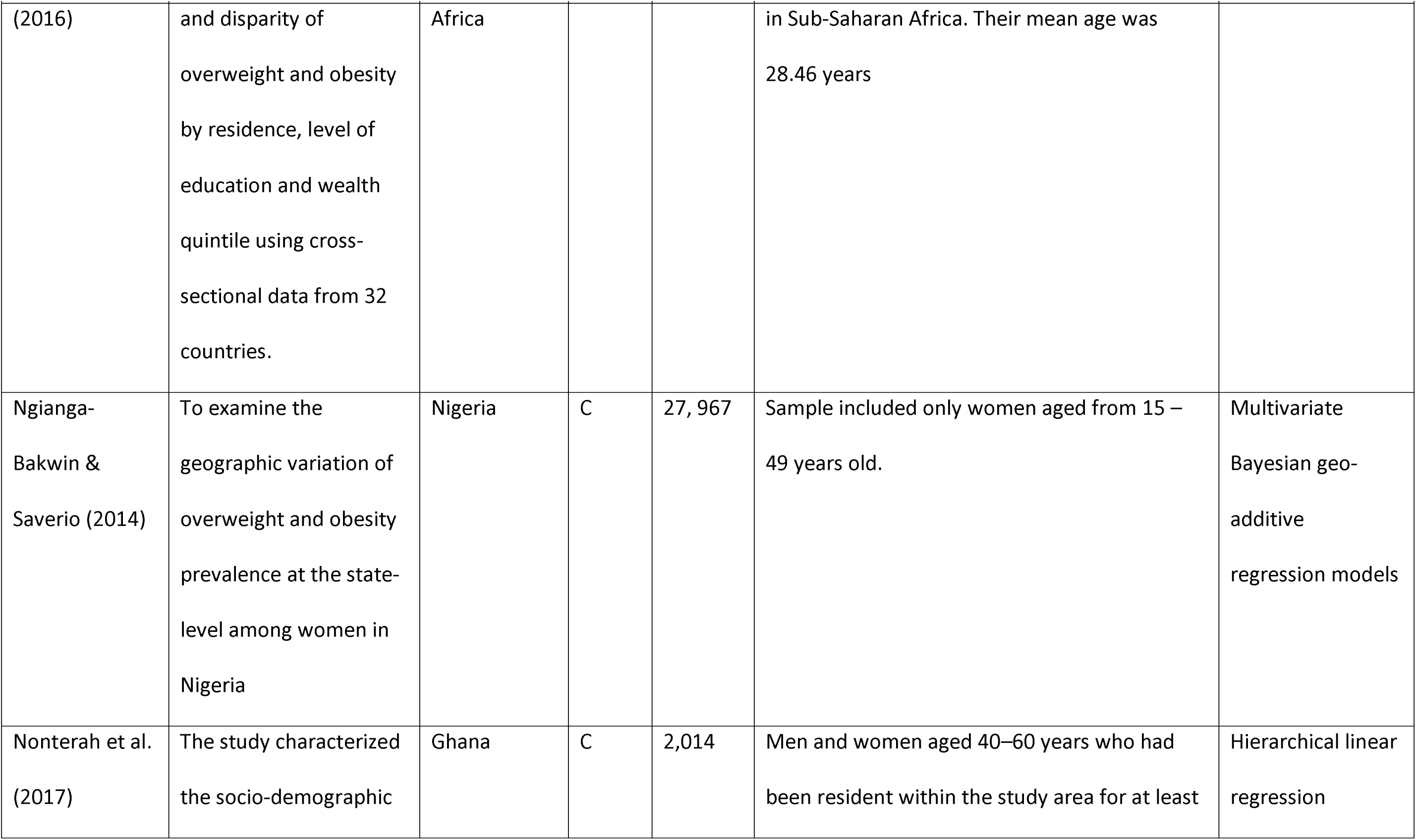
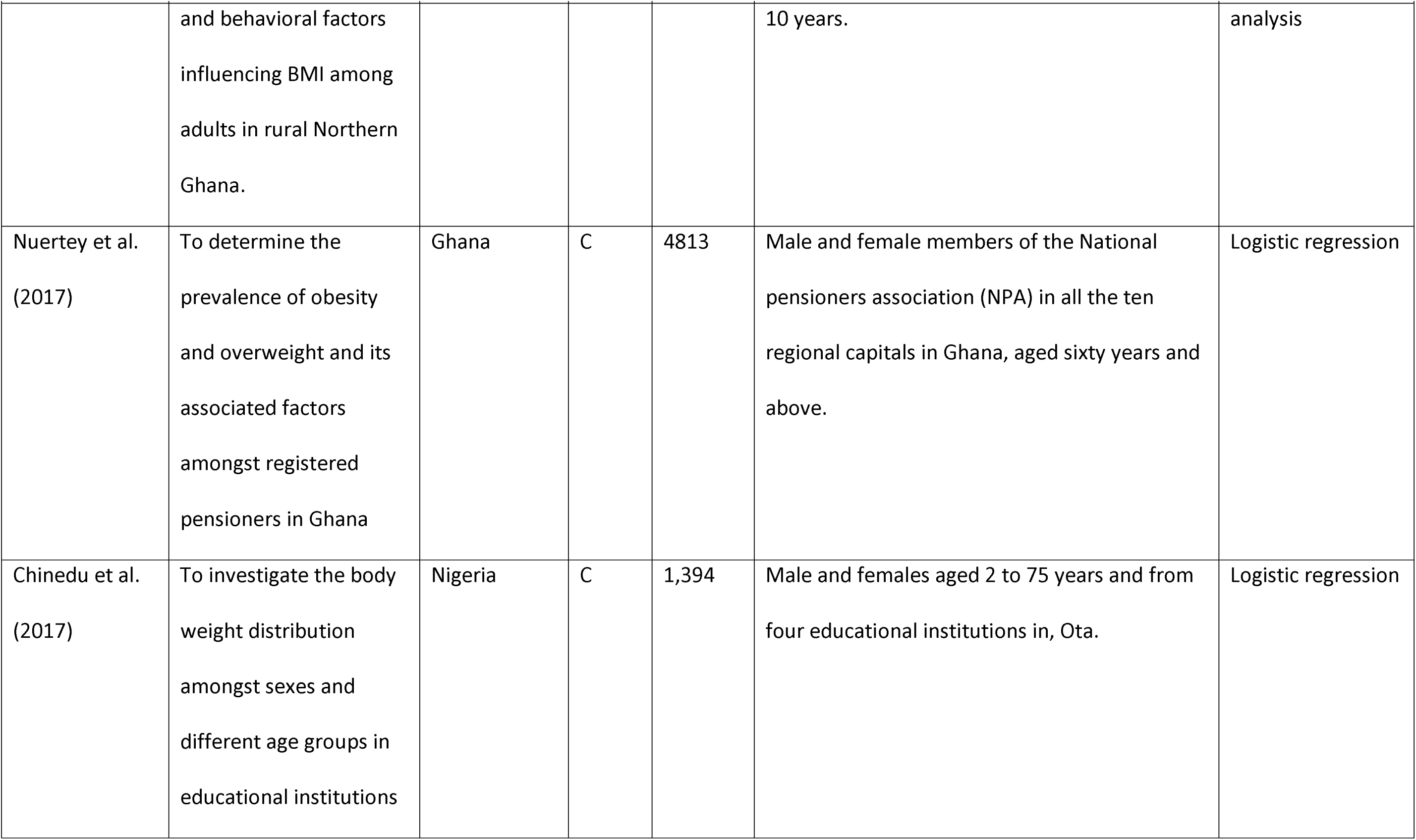
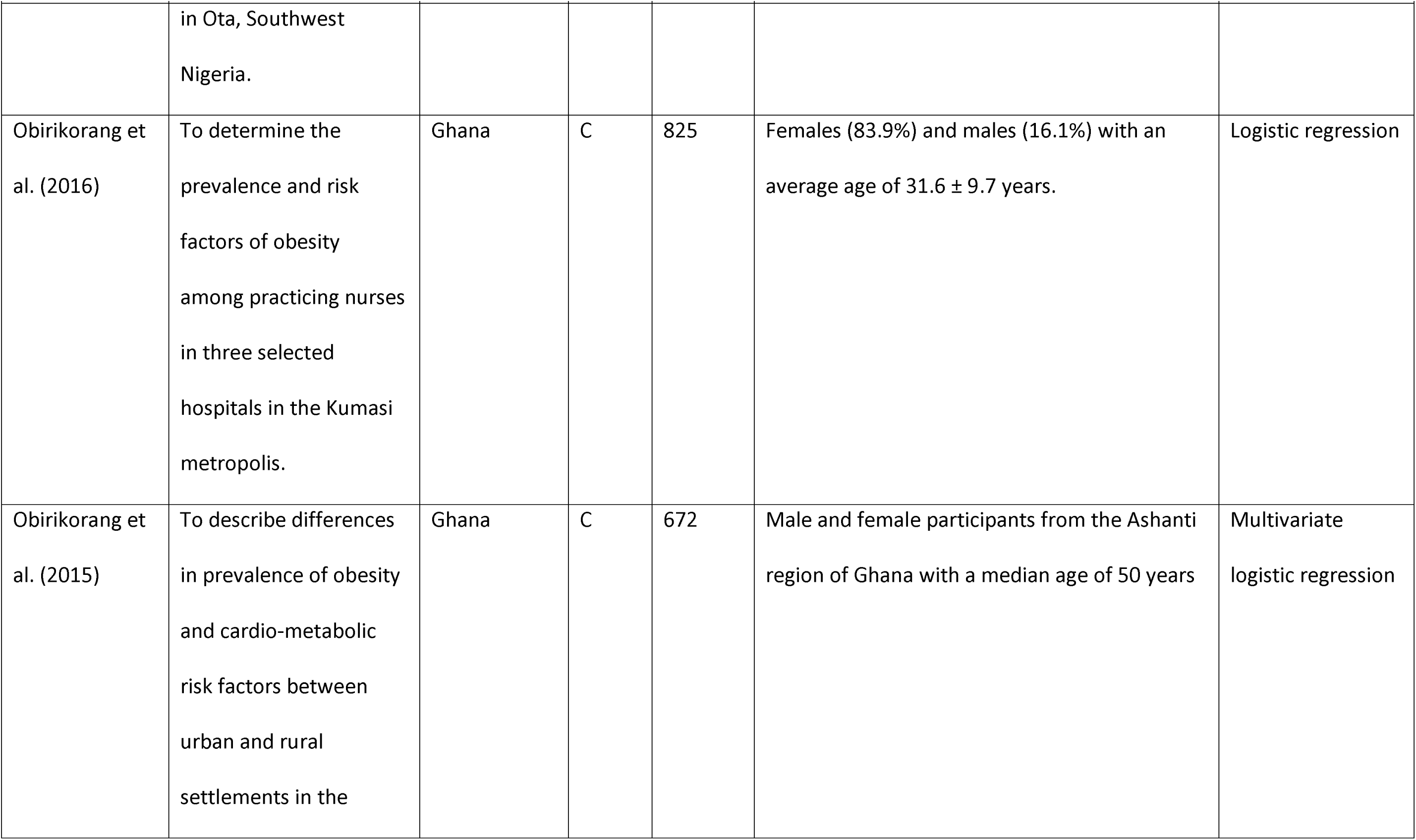
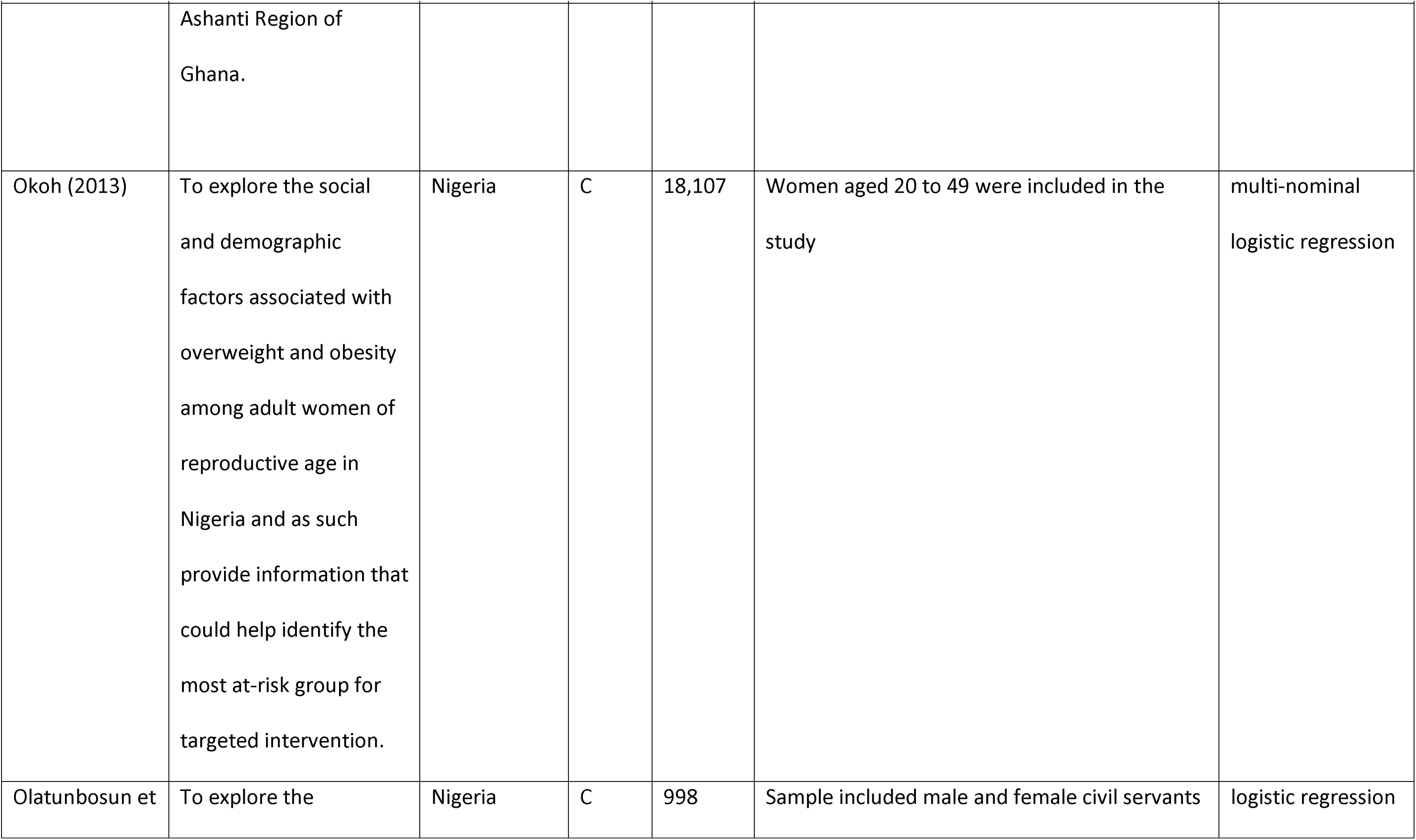
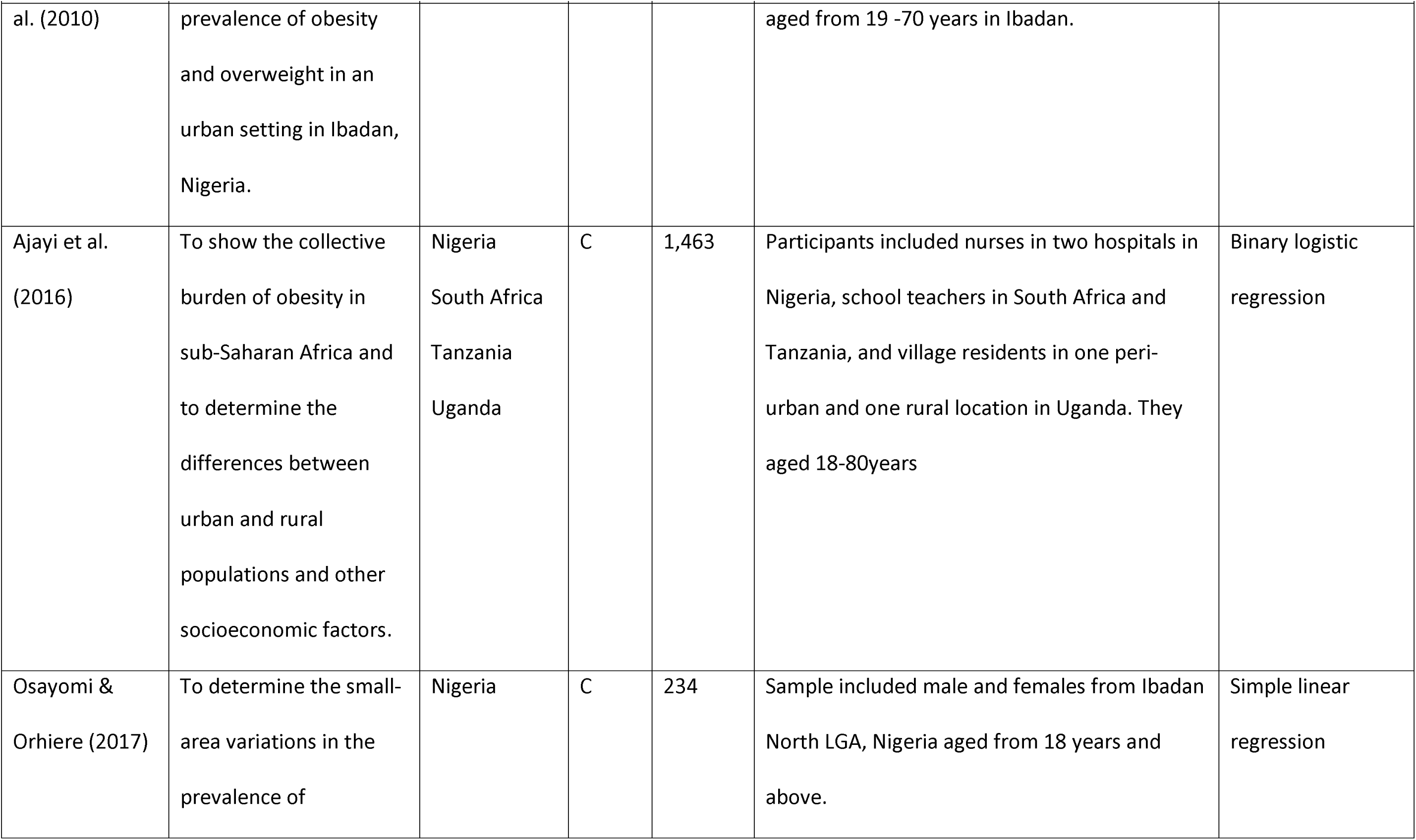
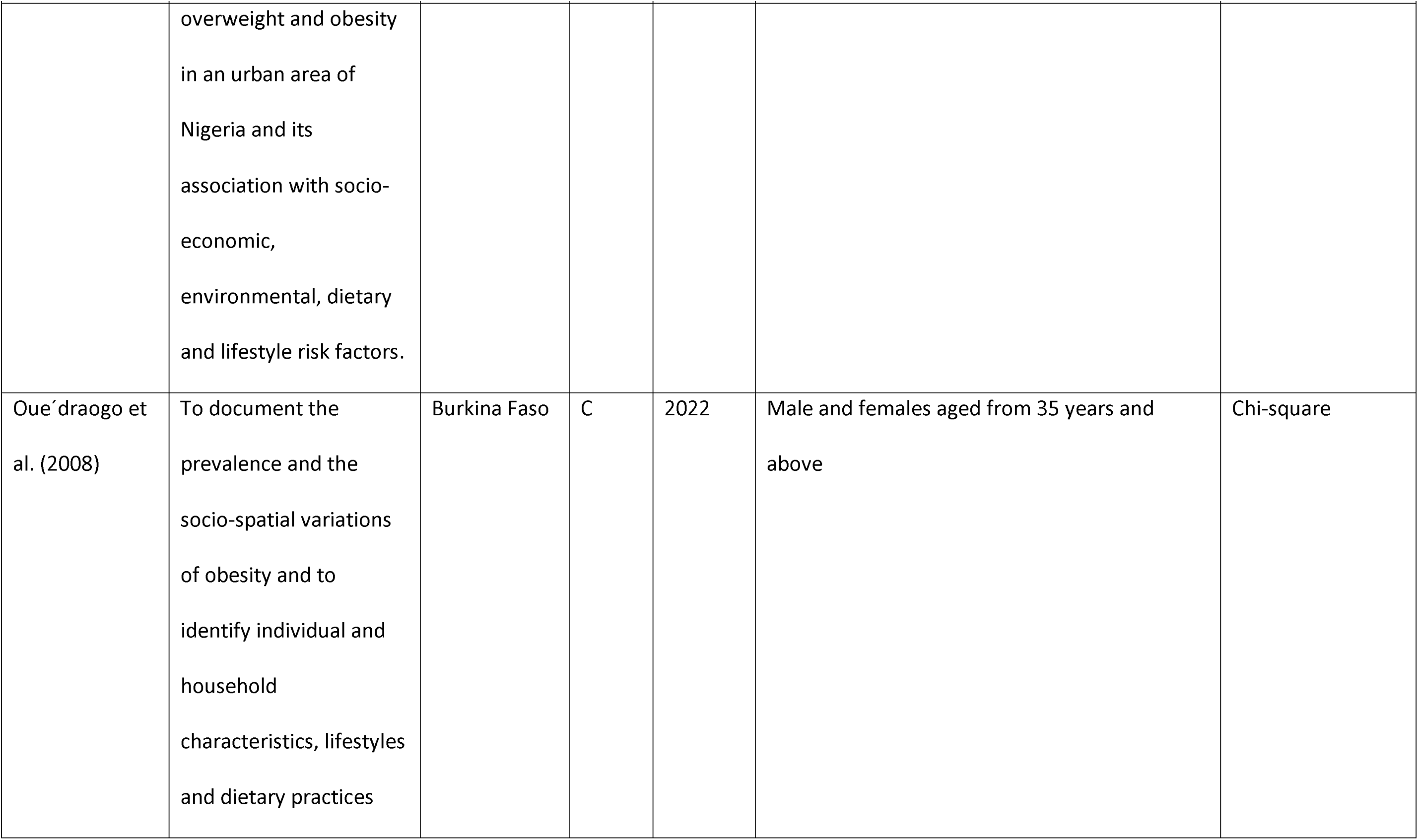
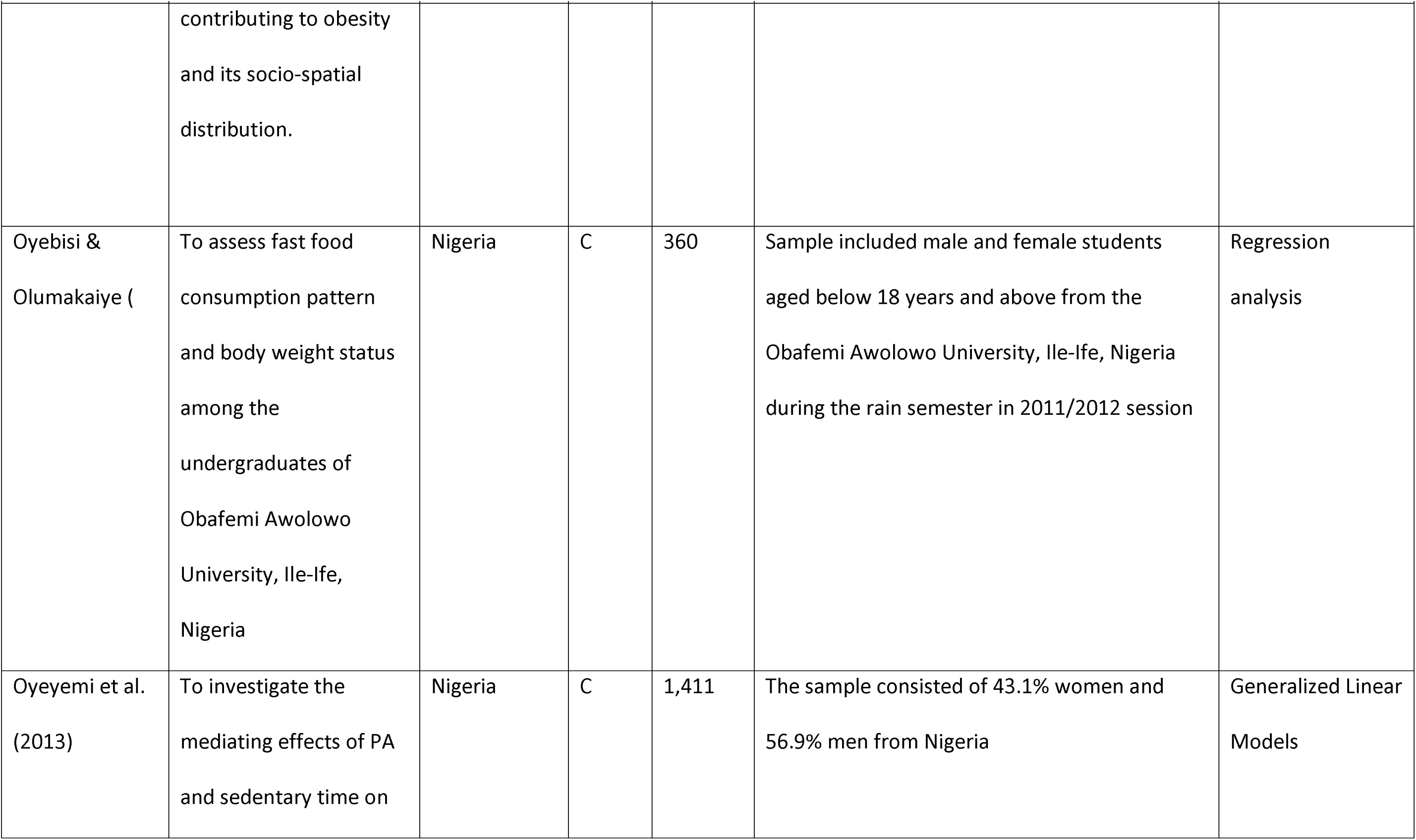
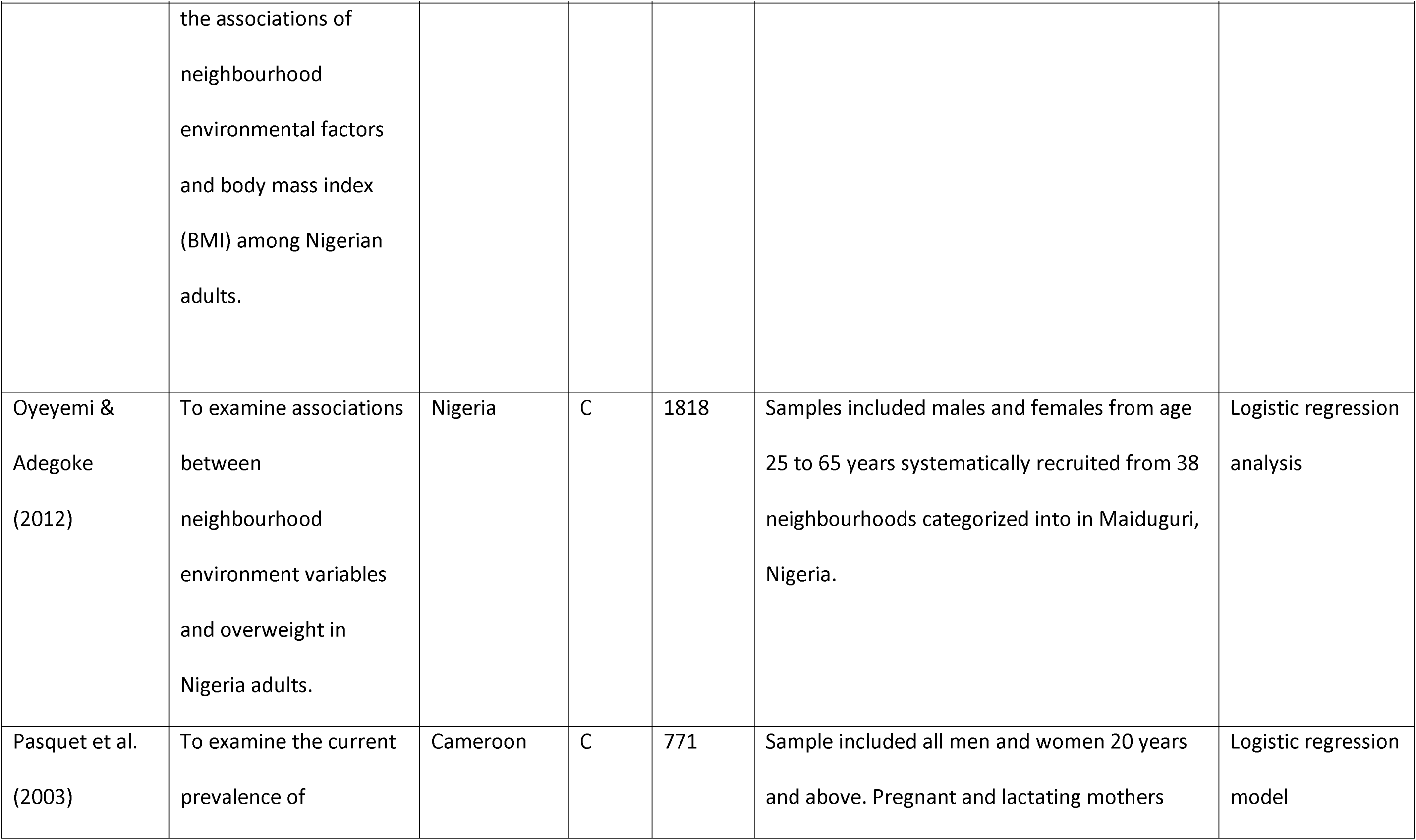
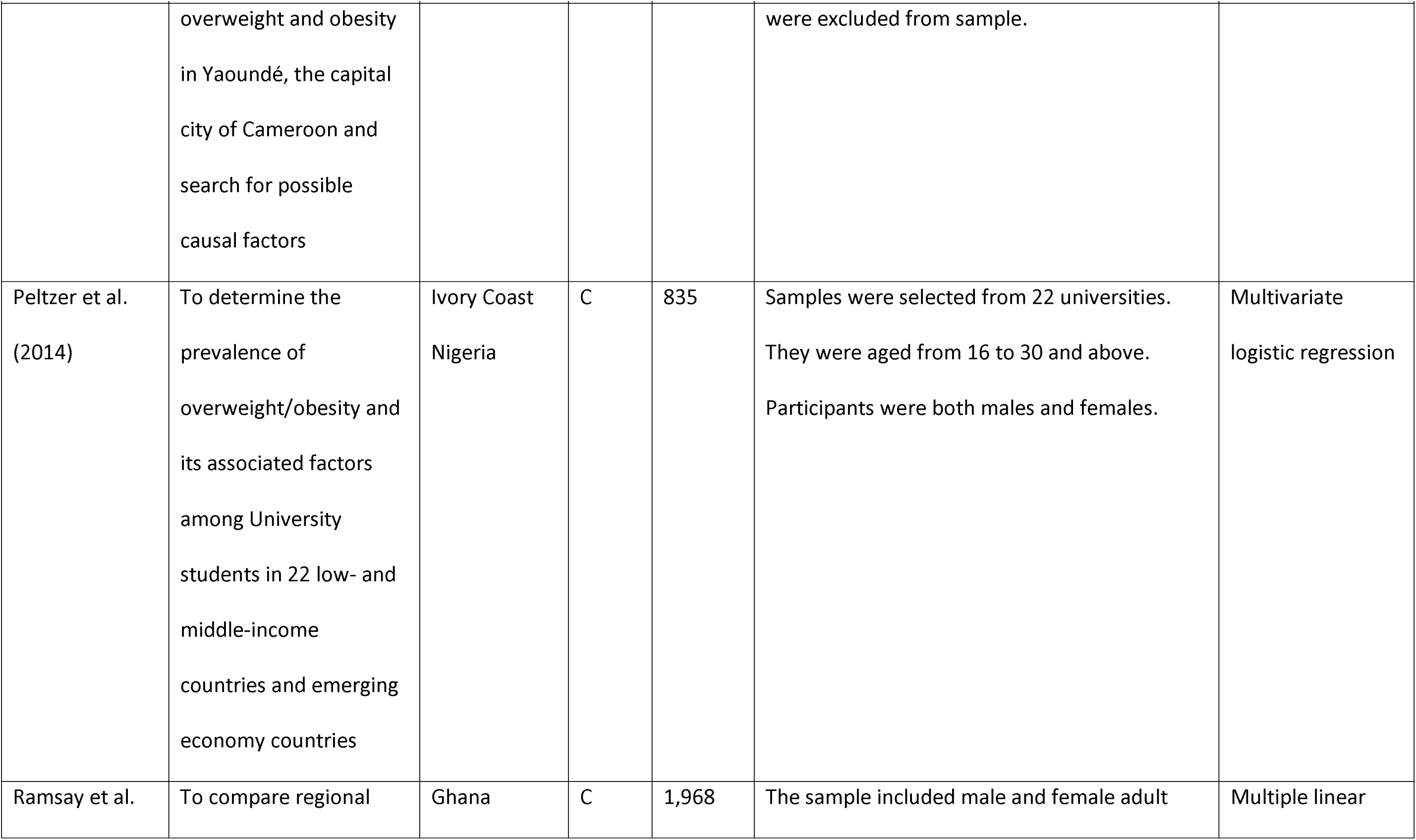
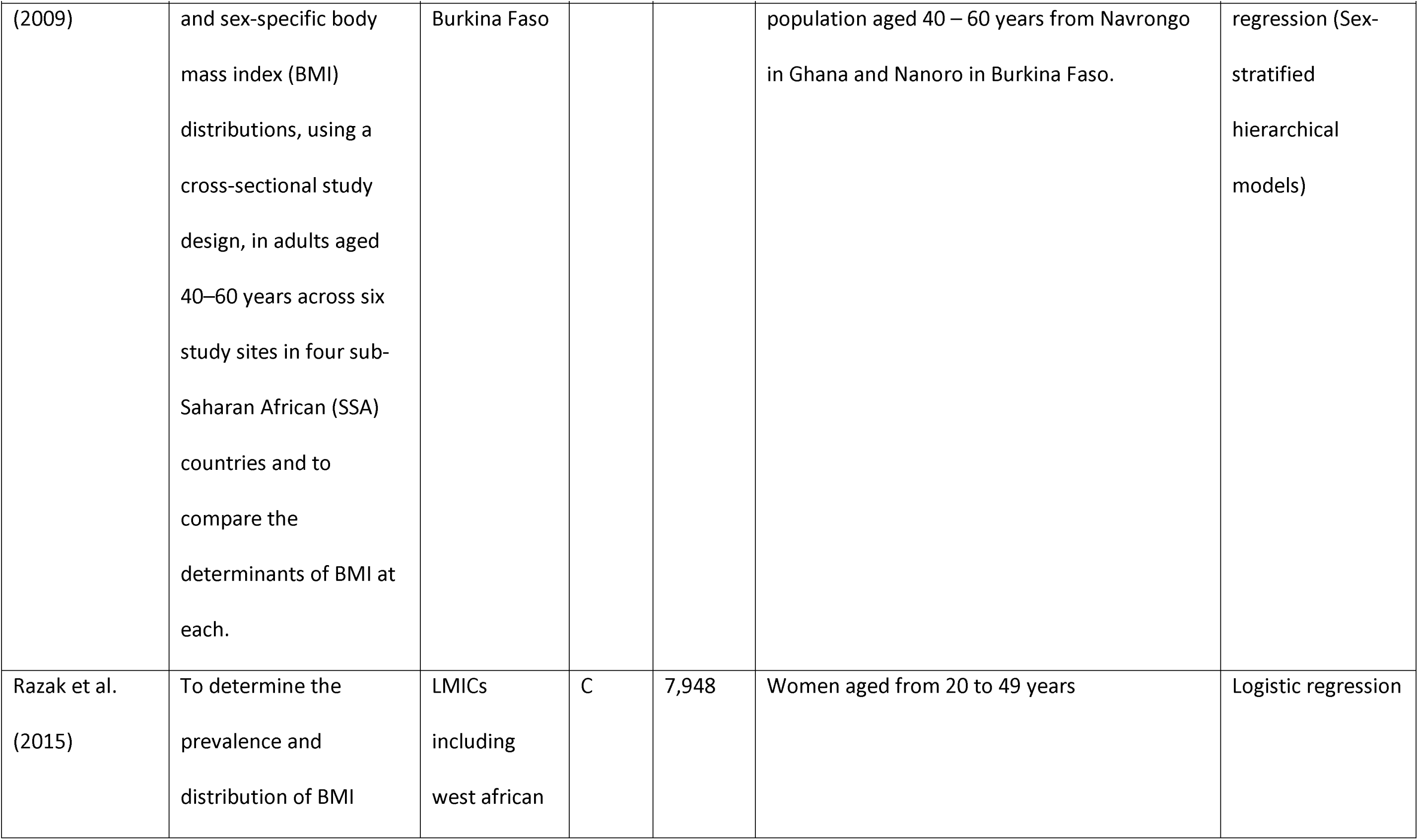
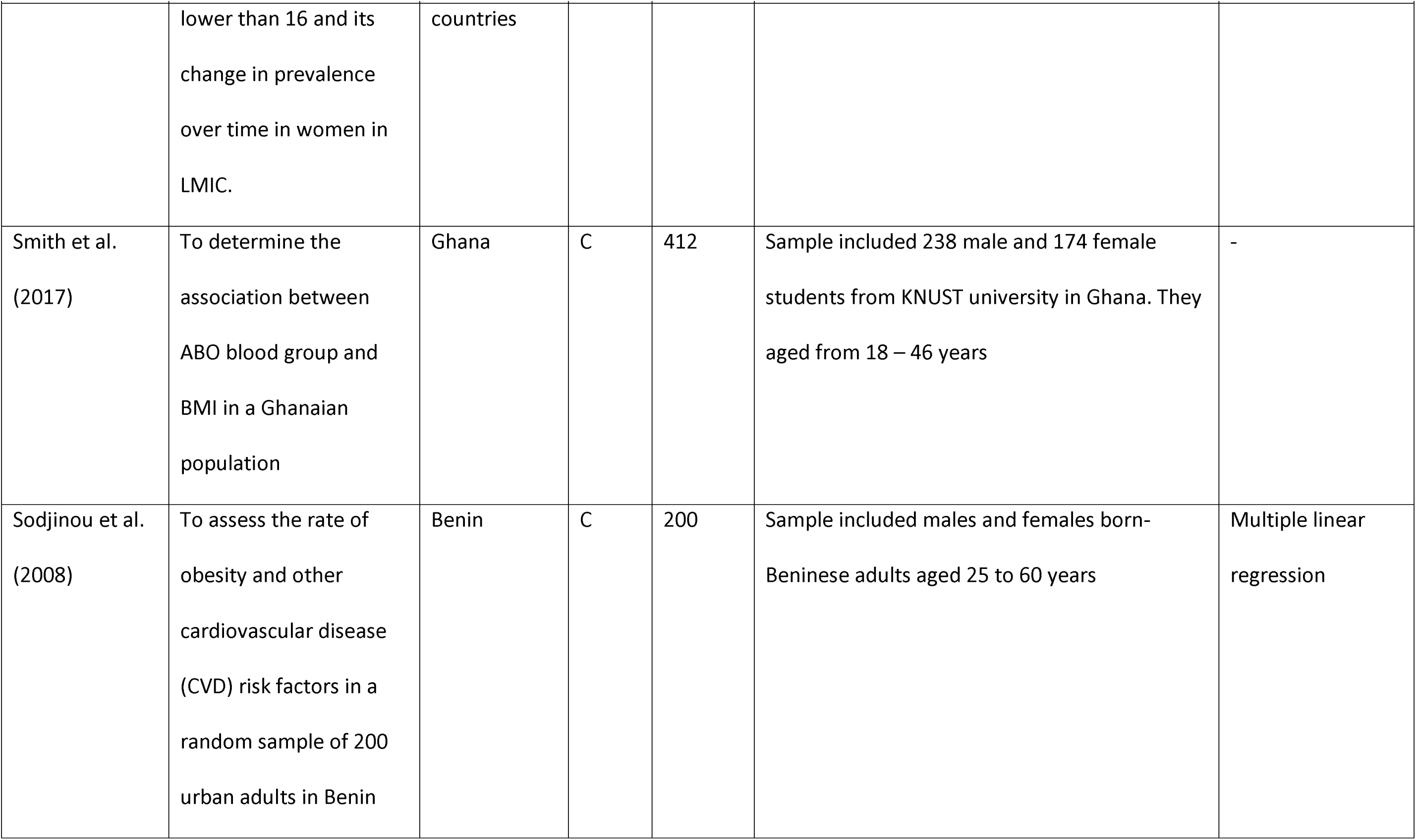
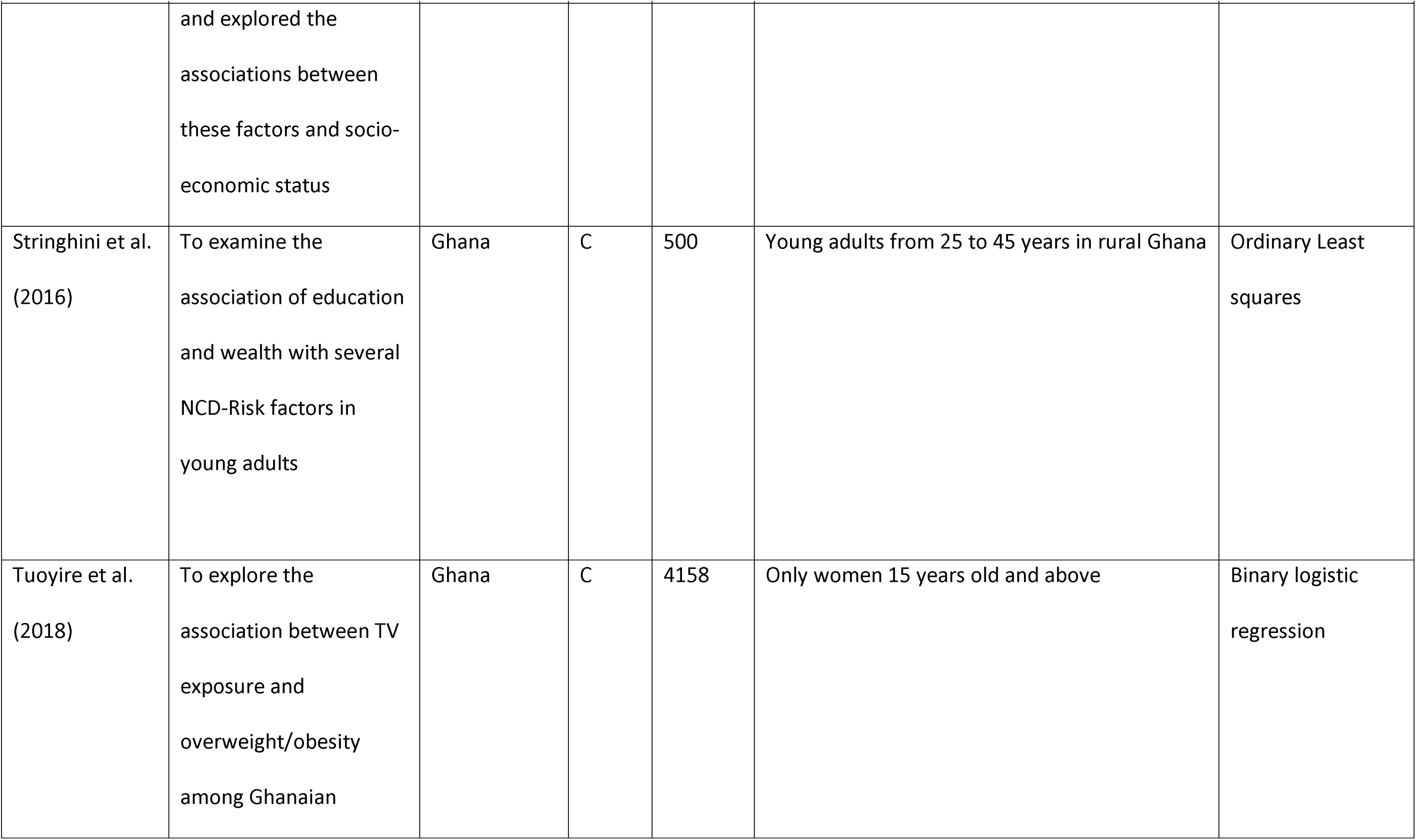
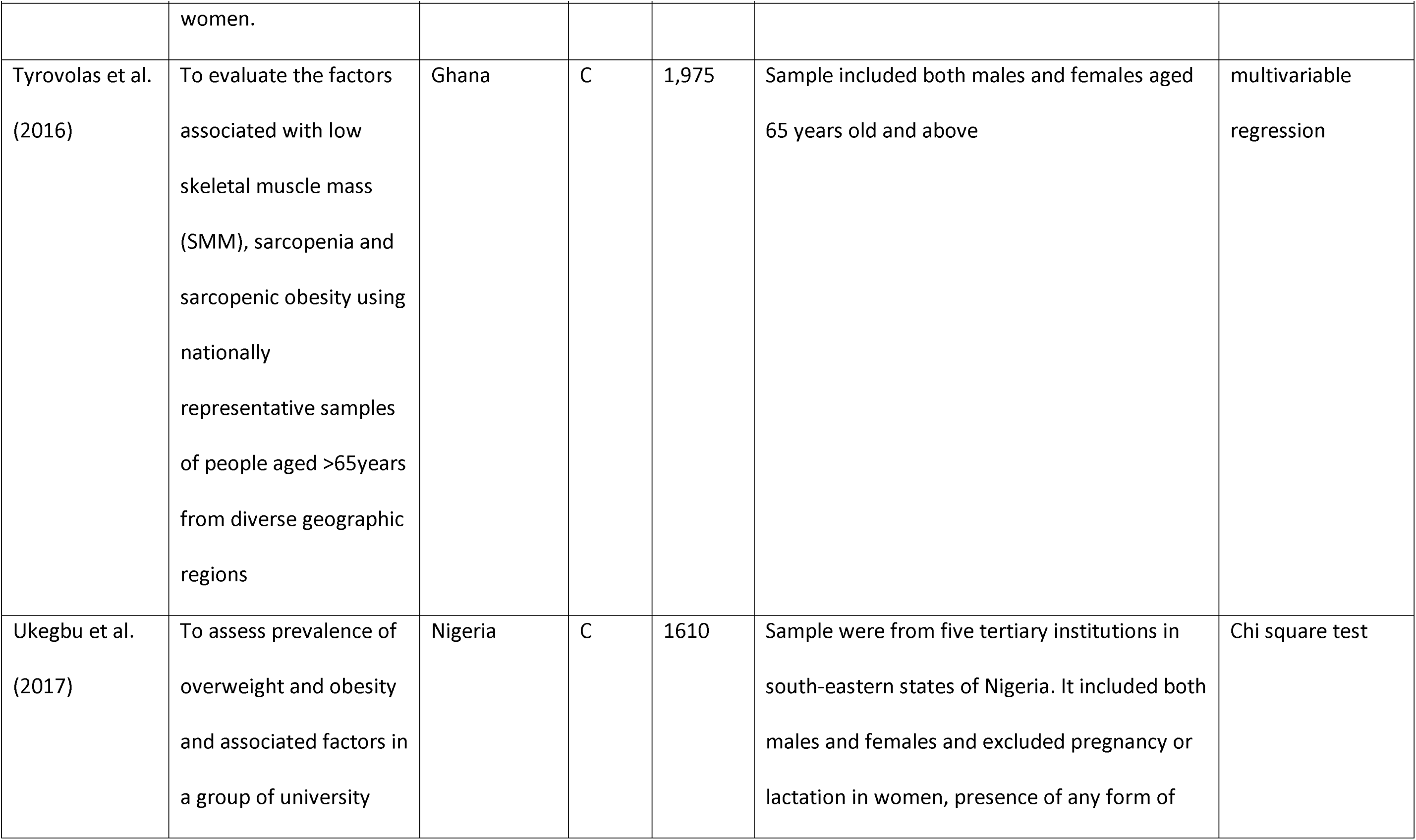
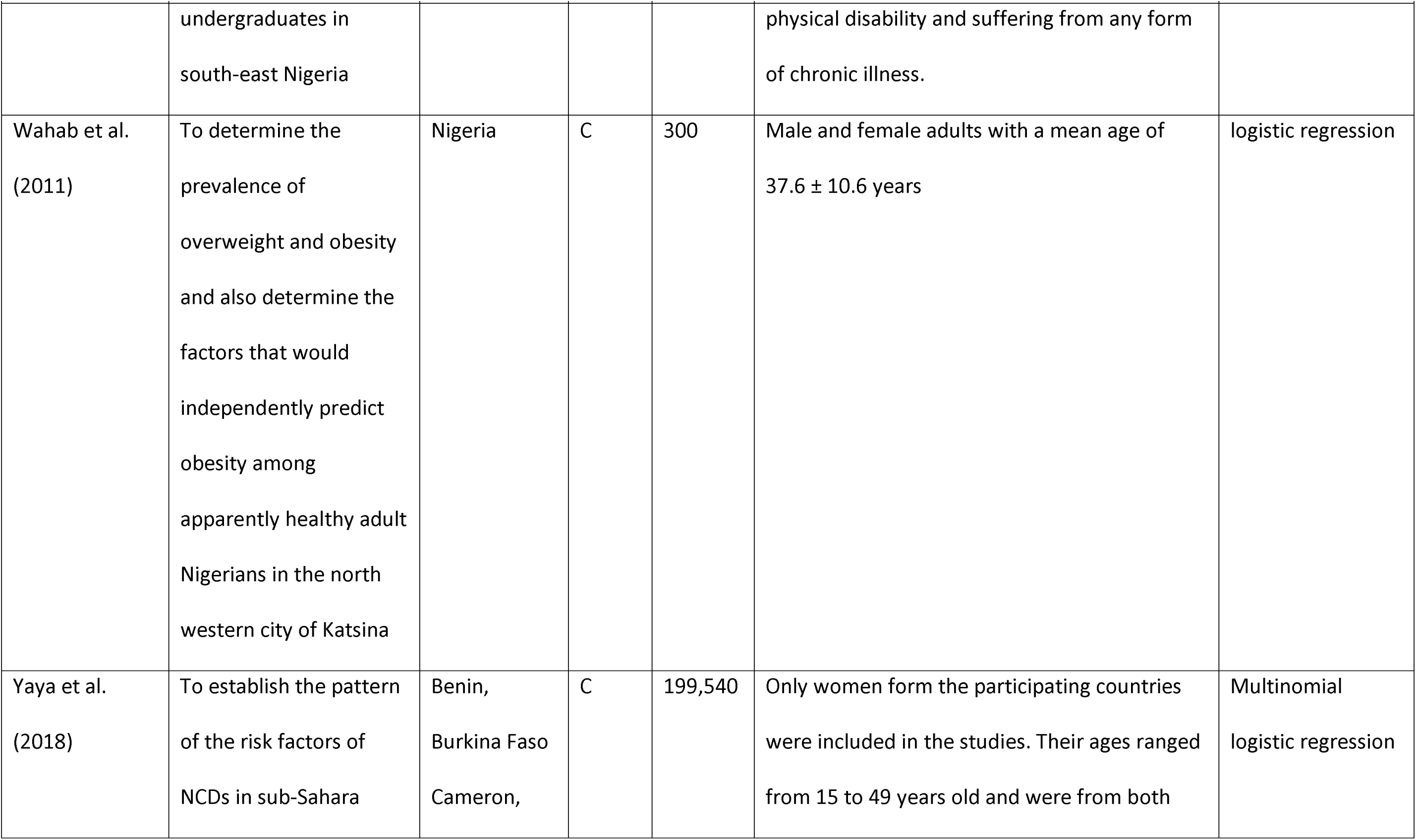
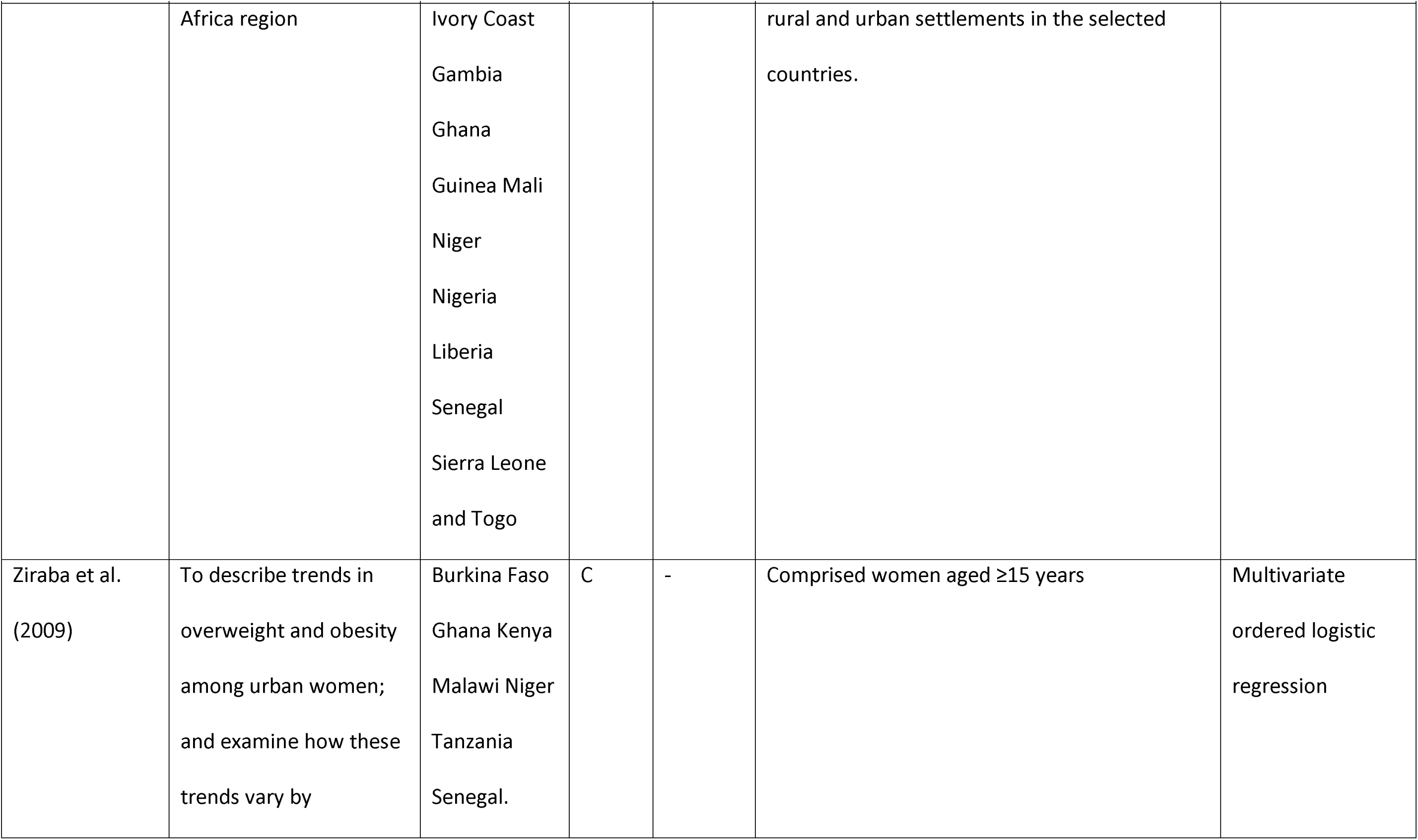
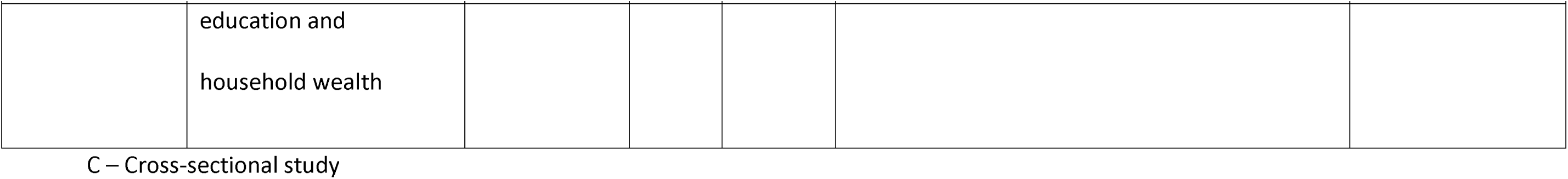
Basic characteristics of the sixty-three included studies

**Table 4:** Independent and dependent variables measured and the study findings

| Author (year) | Determining variables measured | Obesity specification | Findings |
| --- | --- | --- | --- |
| Abdula (2010) | Weight<br>Household size<br>Mother/ Child's age<br>Mother's marriage status<br>Mother's education<br>Ethnicity<br>Mother's occupation<br>Food consumption<br>TV Viewing | WHO BMI measure of obesity;<br>Underweight <18.50 kg/m<sup>2</sup><br>Normal range (18.50–<br>24.99 kg/m<sup>2</sup>)<br>Overweight (25.00–<br>29.99 kg/m<sup>2</sup>)<br>Obese (≥30 kg/m<sup>2</sup>). | Mothers' education, employment status and ethnicity<br>are associated with obesity |
| Addo et al.<br>(2015) | Ages<br>Sex<br>educational level<br>marital status | BMI measure of obesity;<br>Underweight <18.50 kg/m<sup>2</sup><br>Normal range (18.50–<br>24.99 kg/m<sup>2</sup>) | Physical activity (OR = 0.34, 95 % CI = 0.13–0.89,<br>p = 0.03), alcohol consumption (OR = 3.00, 95 %<br>CI = 1.35, 6.68, p = 0.007), marital status (OR = 2.74,<br>95 % CI = 0.96–7.85, p = 0.04), sex (OR = 2.78, 95 % |
|  | Years with institution<br>Sedentary work<br>Alcohol intake | Overweight (25.00–29.99 kg/m<sup>2</sup>)<br>Obese (≥30 kg/m<sup>2</sup>). | CI = 1.23–6.33, p = 0.01), and age (OR = 1.10, 95 % CI = 1.01–1.20, p = 0.036) were associated with obesity and overweight |
| Addo & Leon<br>(2009) | Age<br>Physical activity<br>Level of education<br>Employment grade<br>Current wealth<br>Pre-adult wealth<br>Alcohol consumption<br>Smoking status<br>Ethnicity<br>Place of birth | BMI measure of obesity;<br>Underweight <18.50 kg/m<sup>2</sup><br>Normal range (18.50–24.99 kg/m<sup>2</sup>)<br>Overweight (25.00–29.99 kg/m<sup>2</sup>)<br>Obese (≥30 kg/m<sup>2</sup>). | Positive graded association between pre-adult and adult levels of wealth and the risk of obesity in men (*P* = 0.003), but weak suggestions of an inverse association between adult level of wealth and obesity in women under 45 years of age. |
| Ajayi et al.<br>(2016) | Sex<br>Age<br>Education level | BMI measure of obesity;<br>Underweight <18.50 kg/m<sup>2</sup><br>Normal range (18.50– | Female predicted obesity and overweight in peri-urban Uganda (AOR =8.01, CI =4.02 – 15.96) and obesity in rural Uganda (AOR =11.22, CI =2.27 – 55.40), peri-urban |
|  | Marital status<br>Cigarette smoking<br>Number of cigarettes in past 24h<br>Wealth status<br>Current smoker | 24.99 kg/m<sup>2</sup>)<br>Overweight (25.00–<br>29.99 kg/m<sup>2</sup>)<br>Obese (≥30 kg/m<sup>2</sup>). | Uganda (AOR =27.80, CI =7.13 – 108.41) and South Africa (AOR =2.17, CI: 1.19 – 4.00).<br>Increasing age predicted BMI≥25kg/m<sup>2</sup> in Nigeria (Age>=45, AOR = 9.11, 1.72 – 48.16) and South Africa (AOR =6.22, CI =2.75 – 14.07), while marital status predicted BMI≥ 25kg/m<sup>2</sup> only in peri-urban Uganda (Married – AOR=4.49, CI: 1.74 – 11.57). |
| Akarolo-Anthony et al.<br>(2014) | Age<br>Sex<br>Religion<br>Marital status<br>Education<br>Occupation<br>Socio-economic status<br>Sugar sweetened beverages<br>Physical activity | BMI measure of obesity;<br>Underweight <18.50 kg/m<sup>2</sup><br>Normal range (18.50–<br>24.99 kg/m<sup>2</sup>)<br>Overweight (25.00–<br>29.99 kg/m<sup>2</sup>)<br>Obese (≥30 kg/m<sup>2</sup>). | Compared with the individuals in the lower socio-economic status, the PR of obesity among those in the middle and high socio-economic statuses were 1.39, CI =1.13 – 1.72 and 1.24, CI =0.97 – 1.59 respectively. |
|  | Television |  |  |
| Akinyeniju et al.<br>(2016) | Age<br>Marital status<br>Smoking<br>Alcohol use<br>Health status<br>Education<br>Mother's education<br>Father's education<br>Employment | BMI measure of obesity;<br>Underweight <18.50 kg/m<sup>2</sup><br>Normal range (18.50–<br>24.99 kg/m<sup>2</sup>)<br>Overweight (25.00–<br>29.99 kg/m<sup>2</sup>)<br>Obese (≥30 kg/m<sup>2</sup>). | Participants whose mothers were employed in the public sector had significantly higher BMI compared with those who were unemployed (24.7 vs 22.4); and participants whose fathers were employed in the public sector had higher BMI compared with those whose fathers were unemployed (25.3 vs 19.00). Life-course SES based on both maternal (p=0/0339) and paternal (p= 0.0062) education and paternal employment (p<0.0001) were each associated with BMI |
| Aladeniyi et al.<br>(2017) | Sex<br>Age<br>Level of education<br>Marital status | BMI measure of obesity;<br>Underweight <18.50 kg/m<sup>2</sup><br>Normal range (18.50–<br>24.99 kg/m<sup>2</sup>) | Female (AOR= 5.7 CI =4.7-6.9), age (AOR=1.4, CI = 1.1-1.8), level of education (AOR =0.8, CI=0.7-0.9), marital status (AOR= 2.1, CI= 1.7-2.8), alcohol consumption (AOR= 0.7, CI=0.5-0.9), diabetes mellitus (AOR= 0.7, CI= |
|  | Grade level<br>Excessive alcohol consumption<br>Engaging in physical activity<br>Spending 8 or > hours in sitting<br>Diabetes mellitus<br>High blood pressure | Overweight (25.00–<br>29.99 kg/m<sup>2</sup>)<br>Obese (≥30 kg/m<sup>2</sup>). | 0.5-0.9) and hypertension (AOR=0.5, CI= 0.4-0.6)<br>predict obesity |
| Engle-Stone et al. (2018) | Age<br>Household socio-economic status<br>Exposure to media<br>Intake of fortified food<br>Consumption of processed snack | BMI measure of obesity;<br>Underweight <18.50 kg/m<sup>2</sup><br>Normal range (18.50–<br>24.99 kg/m<sup>2</sup>)<br>Overweight (25.00–<br>29.99 kg/m<sup>2</sup>)<br>Obese (≥30 kg/m<sup>2</sup>). | Household TV ownership (OR= 0.31 CI =0.11, 0.92)] and<br>age (OR =0.38 CI =0.12, 1.20) were associated with<br>obesity |
| Atuahene et al. (2017) | Age<br>Sex<br>Marital status | BMI measure of obesity;<br>Underweight <18.50 kg/m<sup>2</sup><br>Normal range (18.50– | Marital status (p=0.001), leisure time with physical<br>activity (p=0.000) and level of physical activity at work<br>(p=0.035) were significantly associated with BMI. |
|  | Religion<br>Highest education level<br>Alcohol consumption<br>Smoking<br>Most important meal<br>Skipping breakfast<br>Last meal of the day<br>Exercise<br>Leisure time<br>Level of physical activity<br>Means of transport | 24.99 kg/m<sup>2</sup>)<br>Overweight (25.00–<br>29.99 kg/m<sup>2</sup>)<br>Obese (≥30 kg/m<sup>2</sup>). |  |
| Becquey et al.<br>(2010) | Snacking score (frequent food consumption outside the main meals)<br>Modern foods score (modernity of the type of foods consumed) | BMI measure of obesity;<br>Underweight <18.50 kg/m<sup>2</sup><br>Normal range (18.50–<br>24.99 kg/m<sup>2</sup>)<br>Overweight (25.00– | A higher "modern foods" score was associated with a higher prevalence of overweight when confounding factors were accounted for (OR = 1.19 [95% CI 1.03-1.36]) |
| | Age<br>Marital status<br>Religion<br>Ever attended school<br>District | 29.99 kg/m<sup>2</sup>)<br>Obese ($\geq 30$ kg/m<sup>2</sup>). | |
| Ghose B. & Yaya S. (2018) | Frequency of reading newspapers<br>Listening to radio<br>Television (TV) viewing | BMI measure of obesity;<br>Underweight <18.50 kg/m<sup>2</sup><br>Normal range (18.50–<br>24.99 kg/m<sup>2</sup>)<br>Overweight (25.00–<br>29.99 kg/m<sup>2</sup>)<br>Obese ($\geq 30$ kg/m<sup>2</sup>). | Watching TV almost every day and at least once a week were associated with, respectively, 1.6 (CI =1.412 – 1.811) and 1.2 (CI =1.053 – 1.363) times higher odds of being overweight, and 2.7 (CI =2.432 – 3.037) and 1.5 (CI = 1.053 – 1.63) times higher odds of being obese compared with those who never used radio. |
| Blankson & Hall (2010) | Half arm span<br>Mid-upper arm circumference (MUaC)<br>Age | BMI measure of obesity;<br>Underweight <18.50 kg/m<sup>2</sup><br>Normal range (18.50–<br>24.99 kg/m<sup>2</sup>) | BMI was strongly positively correlated with half arm span (r=0.999, P<0.001) and with MUaC (r=0.91, P<0.001). |
|  | Chewing tobacco<br>Drinking alcohol<br>Using walking aid | Overweight (25.00–<br>29.99 kg/m<sup>2</sup>)<br>Obese (≥30 kg/m<sup>2</sup>). |  |
| Boua et al.<br>(2018) | Age<br>Gender<br>Ethnicity<br>Marital status<br>Highest level of education<br>Employment<br>Household asset status<br>Smoking status<br>Snuff use<br>Chewing tobacco<br>Alcohol intake<br>Diet | BMI measure of obesity;<br>Underweight <18.50 kg/m<sup>2</sup><br>Normal range (18.50–<br>24.99 kg/m<sup>2</sup>)<br>Overweight (25.00–<br>29.99 kg/m<sup>2</sup>)<br>Obese (≥30 kg/m<sup>2</sup>). | Age was inversely associated with BMI in men ($\beta$ = -0.09, CI = -0.12 to -0.69). Problematic drinking was associated with a decreased in BMI by 0.89 units (CI = -1.54 to -0.24) compared to those who never consumed alcohol, whereas smoking was found to be associated with a decreased of 2BMI units ($\beta$ = -2.0, CI = -2.59 to -1.41) compared to those who never smoked. Among women, chewing tobacco was associated with a BMI decrease ($\beta$ = -0.79, CI = -1.2 to -0.37). |
|  | Physical activity<br>Clinical history<br>Self-reported diabetes status<br>HIV positive<br>TB positive |  |  |
| Chigbu et al.<br>(2018) | Gender<br>Age<br>Residence<br>Urban class<br>Marital status<br>Education level<br>Income status<br>Ethnicity | BMI measure of obesity;<br>Underweight <18.50 kg/m<sup>2</sup><br>Normal range (18.50–<br>24.99 kg/m<sup>2</sup>)<br>Overweight (25.00–<br>29.99 kg/m<sup>2</sup>)<br>Obese (≥30 kg/m<sup>2</sup>). | Men were less likely to be overweight (AOR 0.79; 95%CI 0.68-0.92) and obese (AOR 0.24; 95%CI 0.19-0.31) than women.<br>Urban residents were more likely to be overweight (AOR 1.42; 95%CI 1.18-1.71) and obese (AOR 2.09; 95%CI 1.58-2.76) than rural residents. Each additional 1-year increase in age increased the risk of overweight by 1.012 (95%CI 1.005-1.018) and obesity by 1.03 (AOR 1.03; 95%CI 1.02-1.04).<br>The low-income class was less likely to be overweight (AOR 0.694; 95%CI 0.507-0.951) and obese (AOR 0.44; |
|  |  |  | 95% CI 0.28-0.67). |
| Chukwuonye et al. (2013) | Hypertension<br>Diabetes mellitus | BMI (Underweight <18.50 kg/m<sup>2</sup><br>Normal range (18.5-24.99 kg/m<sup>2</sup>)<br>Overweight (25.00–29.99 kg/m<sup>2</sup>)<br>Obese (≥30 kg/m<sup>2</sup>).<br>Abdominal obesity (Waist circumference of 102 cm or more in men and 88 cm or more in women | The prevalence of abdominal obesity was higher (21.75%) than the prevalence of obesity based on BMI (11.12%) in the population. |
| Cohen et al. (2018) | Dietary intake<br>Physical activity<br>Health status perception<br>Anthropometry variables | BMI measure of obesity;<br>Underweight <18.50 kg/m<sup>2</sup><br>Normal range (18.50–24.99 kg/m<sup>2</sup>) | Being female, older, living in urban/suburban areas and valuing larger body size were independently associated with being overweight/obese, but not high-calorie diet |
|  | Perceptions of corpulence | Overweight (25.00–<br>29.99 kg/m<sup>2</sup>)<br>Obese (≥30 kg/m<sup>2</sup>). |  |
| Corsi et al.<br>(2010) | Age<br>Education<br>Household wealth<br>Neighbourhood of respondent | BMI measure of obesity;<br>Underweight <18.50 kg/m<sup>2</sup><br>Normal range (18.50–<br>24.99 kg/m<sup>2</sup>)<br>Overweight (25.00–<br>29.99 kg/m<sup>2</sup>)<br>Obese (≥30 kg/m<sup>2</sup>). | Of the total variation in BMI, 17.6% was attributable to countries (SD 2.0, 95%CI 1.7, 2.4) and 10.6% (SD 1.56, 95% CI 1.54, 1.58) was attributable to neighbourhoods in age-adjusted models. Adjusting for individual- and neighbourhood-level covariates reduced the SD attributable to countries and neighbourhoods to 1.9, and 1.17, respectively. Between-country variation was 13.4% (SD 0.75, 95% CI 0.62–0.90) for underweight and 18.9% (SD 0.92, 95% CI 0.76-1.10) for overweight, and between-neighbourhood variation was 7.7% (SD 0.57, 95% CI 0.55-0.58) for underweight and 7.1% (SD 0.56, 95% CI 0.55-0.58) for overweight in the fully-adjusted multinomial model. |
| Dake et al.<br>(2011) | Age<br>place of residence<br>region of residence<br>level of education<br>ethnicity<br>religion<br>occupation<br>marital status<br>parity<br>household wealth quintile | BMI measure of obesity;<br>Underweight <18.50 kg/m<sup>2</sup><br>Normal range (18.50–<br>24.99 kg/m<sup>2</sup>)<br>Overweight (25.00–<br>29.99 kg/m<sup>2</sup>)<br>Obese (≥30 kg/m<sup>2</sup>). | Women who had higher education were about two times more likely to be overweight compared to non-educated women; those from richer households were twice as likely to be overweight and about five times as likely to be obese compared to those from households of average wealth status. The likelihood of a woman being overweight or obese increased with increasing age. |
| Dake et al.<br>(2016) | Characteristics of the food<br>environment<br>Age<br>Sex<br>Marital status<br>Education level | BMI measure of obesity;<br>Underweight <18.50 kg/m<sup>2</sup><br>Normal range (18.50–<br>24.99 kg/m<sup>2</sup>)<br>Overweight (25.00–<br>29.99 kg/m<sup>2</sup>) | The findings showed a 0.2kg/m<sup>2</sup> increase in BMI for every additional convenience store and a 0.1 kg/m<sup>2</sup> reduction in BMI for every out-of-home cooked food place available after controlling for individual socio-demographic characteristics, lifestyle behaviours and community characteristics. |
| | Type of occupation<br>Length of stay in the community<br>Social cohesion<br>Crime level<br>Trust among community members<br>Lifestyle behaviours | Obese ($\geq 30$ kg/m<sup>2</sup>). | |
| Dake (2012) | GPS data (Spatial location, determined using the local spatial autocorrelation (LISA)) | BMI measure of obesity;<br>Underweight <18.50 kg/m<sup>2</sup><br>Normal range (18.50–24.99 kg/m<sup>2</sup>)<br>Overweight (25.00–29.99 kg/m<sup>2</sup>)<br>Obese ($\geq 30$ kg/m<sup>2</sup>). | The results indicate that almost a quarter of clusters in Ghana contain women who are mostly overweight on average. |
| Damorou et al. (2013) | Age<br>Gender | BMI ((Underweight <18.50 kg/m<sup>2</sup> | Low education level (OR 2.45, 95%CI 1.78 – 4.55, p =0.001) and lack of physical activity (OR 3.57, 95%CI |
| | Education level<br>Dietary habits<br>Total cholesterol<br>Fasting blood sugar<br>Blood pressure | Normal range (18.5-24.99 kg/m<sup>2</sup>)<br>Overweight (25.00–29.99 kg/m<sup>2</sup>)<br>Obese ($\geq 30$ kg/m<sup>2</sup>)).<br>and waist circumference (WC): measured as >88cm for females and >102cm for males. | 2.34 – 9.67, p =0.001) were significantly associated with obesity. |
| Dickson et al. (2016) | Maternal DDS<br>Maternal parity<br>Maternal age (in years)<br>Number of household members<br>Number of children under 5 years<br>Education<br>Employment status | BMI: (Underweight <18.50 kg/m<sup>2</sup><br>Normal range (18.5-24.99 kg/m<sup>2</sup>)<br>Overweight (25.00–29.99 kg/m<sup>2</sup>)<br>Obese ($\geq 30$ kg/m<sup>2</sup>)). | Women who consumed an additional unit of DD achieved an increase of 0.245 in BMI for those in the 90th quantile. The effect of household wealth increases for individuals across all quantiles of the BMI distribution. A unit change in the household wealth score was associated with an increase of 0.038 units increase in BMI for individuals in the 5th quantile in |
|  | Household wealth<br>Sex of household head<br>Presence of co-wives |  | Ghana. Also, 0.237 units increased for those in the 90th quantile in Ghana. |
| Diendéré et al.<br>(2019) | Age<br>Marital status Residence<br>Education levels Occupation<br>Total cholesterol<br>Blood pressure<br>Alcohol use<br>Smoking tobacco | BMI: (Underweight<br><18.50 kg/m<sup>2</sup><br>Normal range (18.5-24.99 kg/m<sup>2</sup>)<br>Overweight (25.00–<br>29.99 kg/m<sup>2</sup>)<br>Obese (≥30 kg/m<sup>2</sup>). | Being a resident of a region in the highest urbanization rate quartile, having a high level of total cholesterol (alone or via an interaction with age) and having a high BP was significantly associated with obesity |
| Doku &<br>Neupane (2015) | Age<br>Education<br>Parity<br>Marital status<br>Wealth index | BMI: (Underweight<br><18.50 kg/m<sup>2</sup><br>Normal range (18.5-<br>24.99 kg/m<sup>2</sup>)<br>Overweight (25.00– | Older age was associated with overweight/obesity among both rural (OR=2.21; 95%CI=1.74 -2.82) and urban residents (OR=2.57; 95%CI=2.03 – 3.25).<br>Women with primary (OR=0.44; 95 % CI=0.22 – 0.88) or no education (OR=0.22; 95 % CI=0.11 – |
| | Place of residence<br>Survey year | 29.99 kg/m<sup>2</sup>)<br>Obese ($\geq 30$ kg/m<sup>2</sup>). | 0.44) had lesser likelihood of being overweight/obese, especially among rural residents. Women with no education (OR=0.48; 95 % CI=0.32 – 0.72) were also less likely of being overweight/obese among urban residents. |
| Maruf & Udoji<br>(2015) | Socio-economic status<br>(occupational and educational indices. | BMI: (Underweight<br><18.50 kg/m<sup>2</sup><br>Normal range (18.5-24.99 kg/m<sup>2</sup>)<br>Overweight (25.00–<br>29.99 kg/m<sup>2</sup>)<br>Obese ($\geq 30$ kg/m<sup>2</sup>). | Participants aged 41–60 years (OR 4.29; 95%CI, 3.25–5.67) had higher odds for Obesity and those >60 years (OR 1.72; 95%CI, 1.21–2.43) had higher odds for overweight compared to those aged 18–40 years.<br>Female was associated overweight (OR 1.20; 95% CI, 0.96–1.51) and obesity (OR 2.21; 95% CI, 1.73–2.83).<br>Participants with secondary education had marginally higher odds for overweight (OR 1.15; 95% CI, 0.88–1.51) and obesity (OR 1.17; 95% CI, 0.86–1.59) than those with tertiary education.<br>and while skilled participants had about the same OR for overweight as professionals, their OR for obesity (OR 1.27; 95% CI, 0.67–2.43) was fairly higher than that for professionals. |
| Fezeu at al.<br>(2005) | Age<br>Gender<br>Household amenities quartile<br>Occupation level<br>Alcohol consumption<br>Tobacco smoking<br>Educational level<br>Physical activity quartile | BMI: (Underweight<br><18.50 kg/m<sup>2</sup><br>Normal range (18.5-24.99 kg/m<sup>2</sup>)<br>Overweight (25.00–<br>29.99 kg/m<sup>2</sup>)<br>Obese (≥30 kg/m<sup>2</sup>). | After adjusting for confounding variables, the odds of obesity (3.8, 1.8–7.8), and abdominal obesity (2.2, 1.3–3.6) were significantly higher for men in the high compared with the low occupational level. |
| Fezeu et al.<br>(2008) | Age<br>Alcohol consumption<br>tobacco smoking,<br>educational level. | BMI: (Underweight<br><18.50 kg/m<sup>2</sup><br>Normal range (18.5-<br>24.99 kg/m<sup>2</sup>)<br>Overweight (25.00– | Age-standardized prevalence of BMI ≥25 kg/m<sup>2</sup> increased significantly in the rural area while the age-standardized prevalence of central obesity increased significantly in the urban population (+32% for women and +190% for men) and it persisted after adjustments |
| | | 29.99 kg/m<sup>2</sup>)<br>Obese ($\geq 30$ kg/m<sup>2</sup>).<br>Central obesity: WC $\geq 80$ cm<br>for women and 94 cm for men | for age group, alcohol consumption, tobacco smoking,<br>and level of education. |
| Goryakin et al.<br>(2015) | Social globalization<br>Political globalization<br>Economic globalization<br>Education<br>Occupation<br>Age<br>Number of children | BMI: (Underweight<br><18.50 kg/m<sup>2</sup><br>Normal range (18.5-24.99 kg/m<sup>2</sup>)<br>Overweight (25.00–<br>29.99 kg/m<sup>2</sup>)<br>Obese ($\geq 30$ kg/m<sup>2</sup>). | Economic globalization quartile 2 (r =0.117, p =0.003),<br>economic globalization quartile 3 (r =0.147, p =0.003)<br>and economic globalization quartile 4 (r =0.139, p<br>=0.003) were significantly associated with overweight<br>in women. |
| Grey et al.<br>(2006) | Socio-economic status<br>Educational level<br>Healthy lifestyle score | BMI: (Underweight<br><18.50 kg/m<sup>2</sup><br>Normal range (18.5- | fat-free mass (kg) was<br>significantly higher in males than females |
|  | Western influences | 24.99 kg/m<sup>2</sup>)<br>Overweight (25.00–<br>29.99 kg/m<sup>2</sup>)<br>Obese (≥30 kg/m<sup>2</sup>). |  |
| Koussouh et al.<br>(2019) | Physical activity<br>Blood pressure<br>Education level<br>Socioeconomic level<br>Age<br>Marital status | BMI: (Underweight<br><18.50 kg/m<sup>2</sup><br>Normal range (18.5-24.99 kg/m<sup>2</sup>)<br>Overweight (25.00–<br>29.99 kg/m<sup>2</sup>)<br>Obese (≥30 kg/m<sup>2</sup>). | Age (p=0.006), marital status (p=0.002) and blood pressure (p=0.004) were significantly associated with obesity. |
| Lartey et al.<br>(2019) | Age<br>Sex<br>Education level | BMI: (Underweight<br><18.50 kg/m<sup>2</sup><br>Normal range (18.5- | Not meeting the recommended physical activity level among females was associated with higher odds of obesity (OR = 3.23; 95% CI: 1.13–6.23) and high central |
|  | Marital status<br>Residence/location<br>Household wealth<br>smoking status<br>alcohol consumption<br>Fruit and vegetable intake<br>Physical activity | 24.99 kg/m<sup>2</sup>) Overweight<br>(25.00–29.99 kg/m<sup>2</sup>)<br>Obese (≥30 kg/m<sup>2</sup>).<br>Waist circumference and high<br>central adiposity (determined<br>using sub-Saharan Africa<br>standards | adiposity (OR = 2.19; 95% CI: 1.32–3.63). |
| Luke et al.<br>(2014) | Physical activity<br>Age<br>Gender<br>Employment<br>Education<br>Participant and significant other's<br>occupation<br>Parental education | BMI: (Underweight<br><18.50 kg/m<sup>2</sup><br>Normal range (18.5-24.99 kg/m<sup>2</sup>)<br>Overweight (25.00–<br>29.99 kg/m<sup>2</sup>)<br>Obese (≥30 kg/m<sup>2</sup>). | In the ecological analysis time spent in moderate-to-vigorous activity was inversely associated with BMI ($r^2=-0.71$), and inversely associated with waist circumference in Ghana ($r=-0.15$). |
|  | Household assets and amenities. |  |  |
| Macia et al.<br>(2010) | Age<br>Gender<br>Marital status<br>Education level<br>Ethnicity | BMI: (Underweight<br><18.50 kg/m<sup>2</sup><br>Normal range (18.5-24.99 kg/m<sup>2</sup>)<br>Overweight (25.00–<br>29.99 kg/m<sup>2</sup>)<br>Obese (≥30 kg/m<sup>2</sup>).<br>Waist circumference (WC) ≥80<br>cm for women and 94 cm for<br>men<br>Waist to Hip ratio (WHR) | Ages 30 – 39 (OR 8.02, CI 3.57 – 18.02, P <0.001), 40 –<br>49 (OR =22.51, CI =8.88 – 57.06, P <0.001) and ≥50 (OR<br>=24.4, CI =9.03 – 65.91, P <0.001) were significantly<br>associated with obesity compared to age 20 – 29.<br>Female was also significantly associated with obesity<br>(OR =17.91, CI =9.39 – 34.15, P <0.001) compared to<br>male. |
| Chidozie et al.<br>(2009) | Socioeconomic strata | BMI: (Underweight<br><18.50 kg/m<sup>2</sup><br>Normal range (18.5- | SES was found to be inversely related (p < 0.010) BMI |
|  |  | 24.99 kg/m<sup>2</sup>)<br>Overweight (25.00–29.99 kg/m<sup>2</sup>)<br>Obese (≥30 kg/m<sup>2</sup>). |  |
| Mogre et al.<br>(2015) | Age<br>Sex<br>Food group<br>Physical activity | BMI: (Underweight <18.50 kg/m<sup>2</sup><br>Normal range (18.5–24.99 kg/m<sup>2</sup>)<br>Overweight (25.00–29.99 kg/m<sup>2</sup>)<br>Obese (≥30 kg/m<sup>2</sup>).<br>Waist to Hip Ratio (WHR): 0.9 or less in men and 0.85 or less in women. | BMI was less likely in students who engaged in vigorous physical activity (AOR=0.3, 95%CI=0.1 – 0.7, p=0.004), but more likely in students who consumed fruits and vegetables>=3 days per week (AOR=2.6, 95% CI1.2 – 5.4, p=0.015). Abdominal obesity was also less likely in male students (AOR=0.0, 95% CI 0.0 – 0.5, p=0.017) but more likely in students who consumed roots and tubers>=3 times per week (AOR=8.0, 95 % CI=2.2 – 10.1, p=0.017) and in those who consumed alcoholic and non-alcoholic beverages>=3 times per week (AOR=8.2, 95 % CI=2.2 – 31.1, p=0.002) |
| Neupane et al.<br>(2016) | Place of residence<br>Socio-economic Status<br>Maternal education | BMI: (Underweight<br><18.50 kg/m<sup>2</sup><br>Normal range (18.5-24.99 kg/m<sup>2</sup>)<br>Overweight (25.00–<br>29.99 kg/m<sup>2</sup>)<br>Obese (≥30 kg/m<sup>2</sup>). | The women in urban residence and those who were classified as rich, with respect to the quintile of the wealth index, had higher likelihood of overweight and obesity. In the pooled results, high education was significantly associated with overweight and obesity. |
| Ngianga-Bakwin<br>& Saverio (2014) | Geographic location<br>Age Education level<br>Religion<br>Wealth index Ethnicity<br>Place of residence | BMI: (Underweight<br><18.50 kg/m<sup>2</sup><br>Normal range (18.5-24.99 kg/m<sup>2</sup>)<br>Overweight (25.00–<br>29.99 kg/m<sup>2</sup>)<br>Obese (≥30 kg/m<sup>2</sup>). | Higher education level (OR 1.68 and 95% credible region (CR) 1.38 – 2.00), higher wealth index (OR 3.45, CR 2.98 -4.05), living in urban settings (OR =1.24, CR =1.14 – 1.36) were significantly associated with higher prevalence of obesity. |
| Nonterah et al.<br>(2017) | Age groups in years<br>Ethnicity | BMI: (Underweight<br><18.50 kg/m<sup>2</sup> | Participants with the highest level of education and a high household socio-economic status had higher BMIs |
| | Partnership status<br>Highest level of education<br>Employment status<br>People-to-bedroom density<br>Household SES categories | Normal range (18.5-24.99 kg/m<sup>2</sup>)<br>Overweight (25.00–29.99 kg/m<sup>2</sup>)<br>Obese (≥30 kg/m<sup>2</sup>). | than those in the lowest strata in both men ($\beta = 0.074, p = 0.028$ and $\beta = 0.072, p < 0.001$, respectively) and women ($\beta = 0.174, p = 0.001$ and $\beta = 0.109, p < 0.001$, respectively). Men ($\beta = -0.050; p < 0.001$) and women ($\beta = -0.073; p < 0.001$) of the Nankana ethnic group had a lower BMI than the Kassena ethnic group. Among men, alcohol consumption ($\beta = -0.021; p = 0.001$) and smoking ($\beta = -0.216; p < 0.001$) were associated with lower BMI. Smokeless tobacco was associated with lower BMI among women. Pesticide exposure was associated with higher BMI ($\beta = 0.022; p = 0.022$) among men |
| Nuertey et al. (2017) | Sex<br>Age<br>Marital status | BMI: (Underweight <18.50 kg/m<sup>2</sup><br>Normal range (18.5- | Hypertensive pensioners had 2.6 times odds of being obese compared to non-hypertensive pensioners (95%CI 2.3–2.9). Diabetics were associated with higher |
|  | Ethnicity<br>Education level<br>Social class<br>Region of residence<br>Religion<br>Hypertension<br>Diabetes | 24.99 kg/m<sup>2</sup>)<br>Overweight (25.00–<br>29.99 kg/m<sup>2</sup>)<br>Obese (≥30 kg/m<sup>2</sup>). | odds of being obese (OR 2.5 95% CI 2.0–2.0) compared to non-diabetics. Female pensioners were associated with three times the odds of being obese compared to the male pensioners |
| Chinedu et al.<br>(2017) | Age<br>Sex | BMI: (Underweight<br><18.50 kg/m<sup>2</sup><br>Normal range (18.5-24.99 kg/m<sup>2</sup>)<br>Overweight (25.00–<br>29.99 kg/m<sup>2</sup>)<br>Obese (≥30 kg/m<sup>2</sup>). | Age (r =0.612, p =0.000) was significantly associated with BMI of the subjects. |
| Obirikorang et al. (2016) | Physical activity<br>Alcohol intake | BMI: (Underweight<br><18.50 kg/m<sup>2</sup> | Taking meals late at night (OR 2.5 95%CI 1.1-5.7 p=0.0398) |
|  | Working under stressful hours | Normal range (18.5-24.99 kg/m<sup>2</sup>)<br>Overweight (25.00–29.99 kg/m<sup>2</sup>)<br>Obese (≥30 kg/m<sup>2</sup>). | taking meals at stressful hours [OR=7.9 (2.1 to 29.8); p=0.0009], and fast-food intake [OR=2.6 (1.1 to 6.0), p=0.0370) were independent risk factors of obesity classified by BMI. |
| Obirikorang et al. (2015) | Biochemical Assays<br>measurements<br>Blood pressure<br>Occupation<br>Type of family | BMI: (Underweight <18.50 kg/m<sup>2</sup><br>Normal range (18.5-24.99 kg/m<sup>2</sup>)<br>Overweight (25.00–29.99 kg/m<sup>2</sup>)<br>Obese (≥30 kg/m<sup>2</sup>).<br>Percentage body fat (BF%)<br>estimated using Adult body<br>fat% = (1.20 × BMI) +(0.23 × | Obesity risk factors by urban / rural residence remained significant after adjusting for gender and age |
| | | Age) $-(10.8 \times \text{sex}) -5.4$. | |
| Okoh (2013) | Ethnicity<br>Educational status<br>Marriage status<br>Frequency of watching television | BMI: (Underweight<br>$<18.50 \text{ kg/m}^2$<br>Normal range (18.5-<br>$24.99 \text{ kg/m}^2$)<br>Overweight (25.00–<br>$29.99 \text{ kg/m}^2$)<br>Obese ($\geq 30 \text{ kg/m}^2$). | Results showed increased frequency of watching television, belonging to a particular ethnic group, having a tertiary education and increased parity as risk factors for increased BMI. |
| Olatunbosun et al. (2010) | Age<br>Gender<br>Marital status<br>Salary grade<br>Physical activity<br>Smoking<br>Alcohol<br>Family history of hypertension | BMI: (Underweight<br>$<18.50 \text{ kg/m}^2$<br>Normal range (18.5-<br>$24.99 \text{ kg/m}^2$)<br>Overweight (25.00–<br>$29.99 \text{ kg/m}^2$)<br>Obese ($\geq 30 \text{ kg/m}^2$). | Higher salary grade levels (OR =6.67, $p =0.001$), alcohol use $\geq 60\text{g/week}$ (OR =2.78, $p =0.001$), family history of diabetes (OR =0.38, $p =0.001$) and hypertension (OR =2.68, $p =0.001$) were predictive of obesity and overweight in men. Only age (OR =1.04, $p =0.008$) and higher salary grade levels (OR =4.13, $p =0.002$) were predictive of obesity and overweight for women. |
|  | Family history of diabetes |  |  |
| Ajayi et al.<br>(2016) | Smoking habit<br>Education<br>Gender<br>Age<br>Marital status | BMI: (Underweight<br><18.50 kg/m<sup>2</sup><br>Normal range (18.5-24.99 kg/m<sup>2</sup>)<br>Overweight (25.00–<br>29.99 kg/m<sup>2</sup>)<br>Obese (≥30 kg/m<sup>2</sup>). | Female sex was a predictor of overweight and obesity (combined) in peri-urban Uganda [AOR = 8.01; 95 % CI: 4.02, 15.96) and obesity in rural Uganda [AOR = 11.22; 95%CI: 2.27, 55.40), peri-urban Uganda [AOR = 27.80; 95 % CI: 7.13, 108.41) and SA [AOR = 2. 17; 95 % CI: 1.19, 4.00). Increasing age was a predictor of BMI >=25 kg/m2 in Nigeria [Age >=45 - AOR = 9.11; 95 % CI: 1.72, 48.16] and SA [AOR = 6.22; 95 % CI: 2.75, 14.07], while marital status was predictor of BMI >=25 kg/m2 only in peri-urban Uganda. [Married - AOR = 4.49; 95 % CI: 1.74, 11.57]. |
| Osayomi &<br>Orhiere (2017) | Sex<br>Age<br>Occupation | BMI: (Underweight<br><18.50 kg/m<sup>2</sup><br>Normal range (18.5- | Physical proximity to fast food outlets was the only significant factor driving the spatial pattern of obesity (b = 0.645; R2 = 0.416). |
|  | Education status<br>Ethnicity<br>Marital status<br>Household and neighbourhood characteristics (spatial risk factors) | 24.99 kg/m<sup>2</sup>)<br>Overweight (25.00–<br>29.99 kg/m<sup>2</sup>)<br>Obese (≥30 kg/m<sup>2</sup>). |  |
| Oue'draogo et al. (2008) | Dietary practices<br>Lifestyles | BMI: (Underweight<br><18.50 kg/m<sup>2</sup><br>Normal range (18.5-24.99 kg/m<sup>2</sup>)<br>Overweight (25.00–<br>29.99 kg/m<sup>2</sup>)<br>Obese (≥30 kg/m<sup>2</sup>). | After adjustment for these factors, obesity remained associated with the area of residence: residents from SHBD areas were more likely to be obese than those from ULBD areas (OR=51.41; 95%CI 2.59, 4.76 |
| Oyebisi & Olumakaiye ( | Age<br>Sex<br>Education Base<br>Length of stay in school<br>Upkeep allowance | BMI: (Underweight<br><18.50 kg/m<sup>2</sup><br>Normal range (18.5-24.99 kg/m<sup>2</sup>) | There was a significant relationship between fast food consumption and obesity (r=0.47, p =0.0001). |
|  | Fast food consumption | Overweight (25.00–29.99 kg/m<sup>2</sup>)<br>Obese (≥30 kg/m<sup>2</sup>). |  |
| Oyeyemi et al.<br>(2013) | Physical Activity and Sitting Time<br>Environmental Assessment.<br>Socio-demographic Characteristics | BMI: (Underweight <18.50 kg/m<sup>2</sup><br>Normal range (18.5-24.99 kg/m<sup>2</sup>)<br>Overweight (25.00–29.99 kg/m<sup>2</sup>)<br>Obese (≥30 kg/m<sup>2</sup>). | Walking and total PA significantly mediated the association between BMI and perception of higher residential density (ab ¼_.025 and _.037, respectively), absence of garbage (ab ¼_.046 and _.076, respectively), and more safety from crime at night (ab ¼_.044 and _.083, respectively). In addition, walking, moderate to vigorous PA, and total PA significantly mediated the association between BMI and perception of better aesthetics (ab ¼_.035, _.022, and _.071, respectively). |
| Oyeyemi & Adegoke (2012) | Age, gender, marital status, ethnic group, educational level, employment status, monthly income (naira). | BMI: (Underweight <18.50 kg/m<sup>2</sup><br>Normal range (18.5-24.99 kg/m<sup>2</sup>) | After adjustment for sociodemographic variables, overweight was associated with distant access to commercial facilities (OR 1.49; 95%CI 1.02- 2.18), poor neighbourhood aesthetics (OR1.58; 95%CI 1.16-2.09), |
|  | Residential density, Access to commercial places, Access to non-residential places, Access to public transport, Presence of recreational centres, Presence of pedestrian pathways, Maintenance of pathways, Presence of beautiful things, Absence of unattended animals, Absence of garbage and foul odours, Seeing people active, Connectivity of street, Traffic safety for bicycling, Traffic safety for walking, Crime Safety during the day, Crime safety at night. | Overweight (25.00–29.99 kg/m<sup>2</sup>)<br>Obese (≥30 kg/m<sup>2</sup>). | perceiving garbage and offensive odours in the neighbourhood (OR 1.41 95%CI 1.05-1.89) and feeling unsafe from crime at night (OR 1.47; 95%CI 1.13- 1.91) and unsafe from traffic (OR 1.56; 95%CI 1.17-2.07) in the total sample. Low residential density (OR1.39; 95%CI 1.02-1.93) and poorly maintained pedestrian pathways (OR 1.89; 95% CI 1.13-3.17) associated with overweight in men only, and absence of beautiful things (OR 2.23; 95%CI, 1.42-3.50) and high traffic making it unsafe to walk (OR, 2.39; 95% CI, 1.49-3.83) associated with overweight in women only |
| Pasquet et al.<br>(2003) | Sex<br>Age<br>Education level<br>Occupation length of residence in Yaoundé<br>Ethnicity<br>Parity<br>Smoking practice<br>Physical activity pattern | BMI: (Underweight <18.50 kg/m<sup>2</sup><br>Normal range (18.5-24.99 kg/m<sup>2</sup>)<br>Overweight (25.00–29.99 kg/m<sup>2</sup>)<br>Obese (≥30 kg/m<sup>2</sup>). | The length of residence in Yaoundé (OR =2.0, 95%CI = 1.1 – 3.8) increasing education level (OR =2.7, 95%CI = 1.3 – 5.6), occupation (OR =1.9, 95%CI = 1.1 – 3.6), ethnicity (OR =2.4, 95%CI = 1.3 – 4.4), physical inactivity (OR =1.5, 95%CI = 1.1 – 2.2) and smoking practices (OR =0.5, 95%CI = 0.3 – 0.8) influence early overweight and/or obesity |
| Peltzer et al.<br>(2014) | Age<br>Gender<br>Wealth<br>Country income classification<br>Organized religious activity<br>Social support | BMI: (Underweight <18.50 kg/m<sup>2</sup><br>Normal range (18.5-24.99 kg/m<sup>2</sup>)<br>Overweight (25.00–29.99 kg/m<sup>2</sup>) | Females who are 22years or more are significantly associated with obesity (AOR =1.38, CI: 1.18-1.60).<br>Female in high organized religious activity are significantly associated with overweight and obesity (AOR =1.22, CI =1.03-1.47). |
| | Dietary variables<br>Health risk behaviour | Obese ($\geq 30$ kg/m<sup>2</sup>). | |
| Ramsay et al.<br>(2009) | Age<br>sex<br>ethnicity<br>socio-economic status<br>household crowding<br>education level<br>marital status<br>physical activity<br>sedentary time<br>night-time sleep duration<br>smoking<br>use of smokeless tobacco<br>dietary intake alcohol intake<br>pesticide use<br>HIV and TB status<br>Parity<br>Menopausal stage | BMI: (Underweight<br><18.50 kg/m<sup>2</sup><br>Normal range (18.5-24.99 kg/m<sup>2</sup>)<br>Overweight (25.00–<br>29.99 kg/m<sup>2</sup>)<br>Obese ($\geq 30$ kg/m<sup>2</sup>). | Across Navrongo ($R^2 = 0.20$) and Nanoro ($R^2 = 0.20$),<br>men with higher socioeconomic and educational level<br>were associated with higher BMI. |
| Razak et al.<br>(2015) | Demographic variables<br>Socioeconomic variables | BMI: (Underweight<br><18.50 kg/m<sup>2</sup><br>Normal range (18.5-24.99 kg/m<sup>2</sup>)<br>Overweight (25.00–<br>29.99 kg/m<sup>2</sup>)<br>Obese (≥30 kg/m<sup>2</sup>). | Increased risk of mortality among those with a BMI<br>lower than 16kg/m<sup>2</sup> exceeds the increased risk<br>associated with being overweight or obesity. |
| Smith et al.<br>(2017) | Age<br>Gender<br>ABO Blood groups | BMI: (Underweight<br><18.50 kg/m<sup>2</sup><br>Normal range (18.5- | No significant associations were found between the<br>determining variables and obesity |
|  | Rhesus factor | 24.99 kg/m<sup>2</sup>)<br>Overweight (25.00–<br>29.99 kg/m<sup>2</sup>)<br>Obese (≥30 kg/m<sup>2</sup>). |  |
| Sodjinou et al.<br>(2008) | Diet quality score<br>Alcohol consumption score<br>Smoking score<br>Household amenities<br>Physical activity score | BMI: (Underweight<br><18.50 kg/m<sup>2</sup><br>Normal range (18.5-24.99 kg/m<sup>2</sup>)<br>Overweight (25.00–<br>29.99 kg/m<sup>2</sup>)<br>Obese (≥30 kg/m<sup>2</sup>).<br>Abdominal obesity | After controlling for age and sex, the odds of obesity increased significantly with SES.<br>The likelihood of obesity decreased significantly as while controlling for potential confounding factors. |
| Stringhini et al.<br>(2016) | Smoking<br>Physical activity | BMI: (Underweight<br><18.50 kg/m<sup>2</sup> | Association between SES and obesity was direct |
|  | Weight<br>Height<br>Hypertension<br>Socio-economic indicators | Normal range (18.5-24.99 kg/m<sup>2</sup>)<br>Overweight (25.00–29.99 kg/m<sup>2</sup>)<br>Obese (≥30 kg/m<sup>2</sup>). |  |
| Tuoyire et al.<br>(2018) | Presence of TV<br>Frequency of viewing TV<br>Demographic variables | BMI: (Underweight <18.50 kg/m<sup>2</sup><br>Normal range (18.5-24.99 kg/m<sup>2</sup>)<br>Overweight (25.00–29.99 kg/m<sup>2</sup>)<br>Obese (≥30 kg/m<sup>2</sup>). | Women with TV in their households, and with high TV exposure were significantly (P < 0.05) more likely (OR = 1.39, 95% CI = 1.002, 1.923) to be overweight/obese compared to those with no TV in their households, and no TV exposure after controlling for confounders |
| Tyrovolas et al.<br>(2016) | Age<br>Sex<br>Education | BMI: (Underweight <18.50 kg/m<sup>2</sup><br>Normal range (18.5- | Compared to high levels of physical activity, low levels were related with higher odds for sarcopenia (OR =1.36, 95%CI= 1.11 – 1.67) and sarcopenic obesity (OR |
|  | Wealth<br>Current drinker<br>Current smoker<br>Physical activity<br>Number of chronic conditions | 24.99 kg/m<sup>2</sup>)<br>Overweight (25.00–<br>29.99 kg/m<sup>2</sup>)<br>Obese (≥30 kg/m<sup>2</sup>). | =1.80, CI =1.23 – 2.64) in the overall sample. |
| Ukegbu et al.<br>(2017) | Age<br>Gender<br>Level in school<br>Monthly allowance<br>Number of meals<br>Consumption of unhealthy snack<br>foods<br>Soft drink consumption<br>Weekly alcohol consumption<br>Hypertension | BMI: (Underweight<br><18.50 kg/m<sup>2</sup><br>Normal range (18.5-24.99 kg/m<sup>2</sup>)<br>Overweight (25.00–<br>29.99 kg/m<sup>2</sup>)<br>Obese (≥30 kg/m<sup>2</sup>). | Consumption of unhealthy snack foods ($X^2$ =13.39, $p$=0.037), being female ($X^2$ =47.91, $p$ =0.000), first year student ($X^2$ =41.82, $p$=0.000) and having high systolic ($X^2$ =88.18, $p$=0.000) and diastolic ($X^2$ =10.17, $p$=0.000) pressure were associated with obesity |
| Wahab et al.<br>(2011) | age,<br>sex,<br>current history of alcohol or tobacco use,<br>hypertension and<br>diabetes mellitus | BMI: (Underweight<br><18.50 kg/m<sup>2</sup><br>Normal range (18.5-24.99 kg/m<sup>2</sup>)<br>Overweight (25.00–<br>29.99 kg/m<sup>2</sup>)<br>Obese (≥30 kg/m<sup>2</sup>). | female sex (OR 6.119, 95% CI 2.705-13.842, p < 0.001),<br>hypercholesterolaemia (OR 2.138, 95% CI 1.109- 4.119,<br>p = 0.023) and hyperuricaemia (OR 2.906, 95% CI 1.444-5.847, p = 0.003) were significantly associated with<br>obesity |
| Yaya et al.<br>(2018) | Age<br>Residence<br>Educational level<br>Religion<br>Marital status<br>Wealth index<br>Working<br>Number of children | BMI: (Underweight<br><18.50 kg/m<sup>2</sup><br>Normal range (18.5-24.99 kg/m<sup>2</sup>)<br>Overweight (25.00–<br>29.99 kg/m<sup>2</sup>)<br>Obese (≥30 kg/m<sup>2</sup>). | Rural women were 1.25 times as likely to be<br>underweight, compared with urban women<br>(RR=1.25; 95%CI=1.22–1.28); rural dwellers had<br>51% (RR=0.49; 95%CI=0.48–0.50) and 72%<br>(RR=0.28; 95%CI=0.27–0.29) reduction in<br>overweight and obesity relative to women with normal<br>BMI compared with women from urban residence.<br>Married women or those living with partners had 35% |
|  | Alcohol consumption<br>Currently smoking<br>Exercise<br>Fruit consumption<br>Vegetable consumption |  | reduction in underweight, compared with those never married (RR=0.65; 95%CI=0.64–0.67); however, married women or those currently living with partners were 1.81 (RR=1.81; 95%CI=1.76–1.86) and 2.91 (RR=2.91; 95%CI=2.77–3.04) as likely to be overweight and obesity, compared with never married women.<br>Smoking increased the risk of underweight (RR=2.03; 95%CI=1.92–2.16), but reduced the risk of overweight (RR=0.70; 95%CI=0.65–0.75) and obesity (RR=0.78; 95%CI=0.70–0.86) |
| Ziraba et al.<br>(2009) | Wealth index/ Amenities<br>Time (in years)<br>Woman's education | BMI: (Underweight<br><18.50 kg/m<sup>2</sup><br>Normal range (18.5-<br>24.99 kg/m<sup>2</sup>)<br>Overweight (25.00– | The odds ratio of the variable *time lapse* was 1.05 (p < 0.01), indicating that the prevalence of overweight/obesity increased by about 5% per year on average in the countries in the study. |
|  |  | 29.99 kg/m<sup>2</sup>)<br>Obese (≥30 kg/m<sup>2</sup>). |  |

### Empirical findings on the determinants of obesity in West Africa

Sex was one of the common demographic variables identified in the included studies. Eleven out of the studies^19, 20, 21, 22, 23, 24, 25, 26, 27, 28, 29^ found women to be a significant determinant of obesity when the influence of other variables like age, marital status, education level and employment status are controlled. These studies were from Ghana (n=3), Nigeria (n=5), Nigeria & Ivory Coast (combined) (n=1) and Senegal (n=2). Increasing age was also found as a demographic determinant of obesity in this review^19, 20, 24, 25, 26, 27, 30, 31, 32^ across Ghana (n=2), Nigeria (n=5), Burkina Faso (n=1), Senegal (n=1). In terms of biological factors, only two studies^19, 33^ found hypertension and diabetes as positive determinants of obesity. These findings were from Ghana and Nigeria.

Regarding lifestyle variables, four of the studies, one each from Ghana^36^, Burkina Faso^30^ and Cameroon^34^, and one from thirteen combined countries, namely: Benin, Burkina Faso, Cameron, Ivory Coast, Gambia, Ghana, Guinea, Mali, Niger, Nigeria, Liberia, Senegal, Sierra Leone and Togo^35^, found cigarette smoking to be significantly associated with obesity. Similarly, four of the studies from Ghana^25, 36^, Nigeria^19^ and Burkina Faso^30^ found alcohol consumption as a determinant of obesity. Also, ten studies Nigeria^19, 40^ (n=2), Ghana^23, 25, 28, 37, 38, 41^ (n=6), Togo^39^ (n=1) and Cameroon^34^ (n=1) indicated that lack and low levels of physical activity showed that physical activity is significantly associated with obesity. Additionally, ^42, 28^from Nigeria and Ghana found poor dietary habits as determinants of obesity. All these lifestyle variables were identified as determinants after the studies had controlled for other measured variables.

The socio-economic variables that were identified in the selected papers as determinants of obesity are education level (n=9), employment status (n=3) and Socio-economic status (n=3). These studies were also from Ghana (n=2), Nigeria (n=4), Togo (1), Burkina Faso (n=1) and Cameroon (n=1). Of the environmental factors, most of the variables and the subsequent findings were heterogeneous across all the six studies that measured environmental factors. For instance, Dake et al. (2016)’s^43^ study on the influence of local food environment in a poor urban setting on obesity discovered that for every additional convenience store in poor urban settings in Ghana, there was a 0.2kg/m2 increase in BMI, and every out-of-home cooked food place available, there was a 0.1kg/m2 reduction in BMI of residents while the study by Osayomi and Orhiere (2017)^43^ on environmental factors and obesity in Nigeria identified that physical proximity to fast food outlets is a determinant of obesity. See tables 4 and 5 for a summary of the review.

**Table 5:**
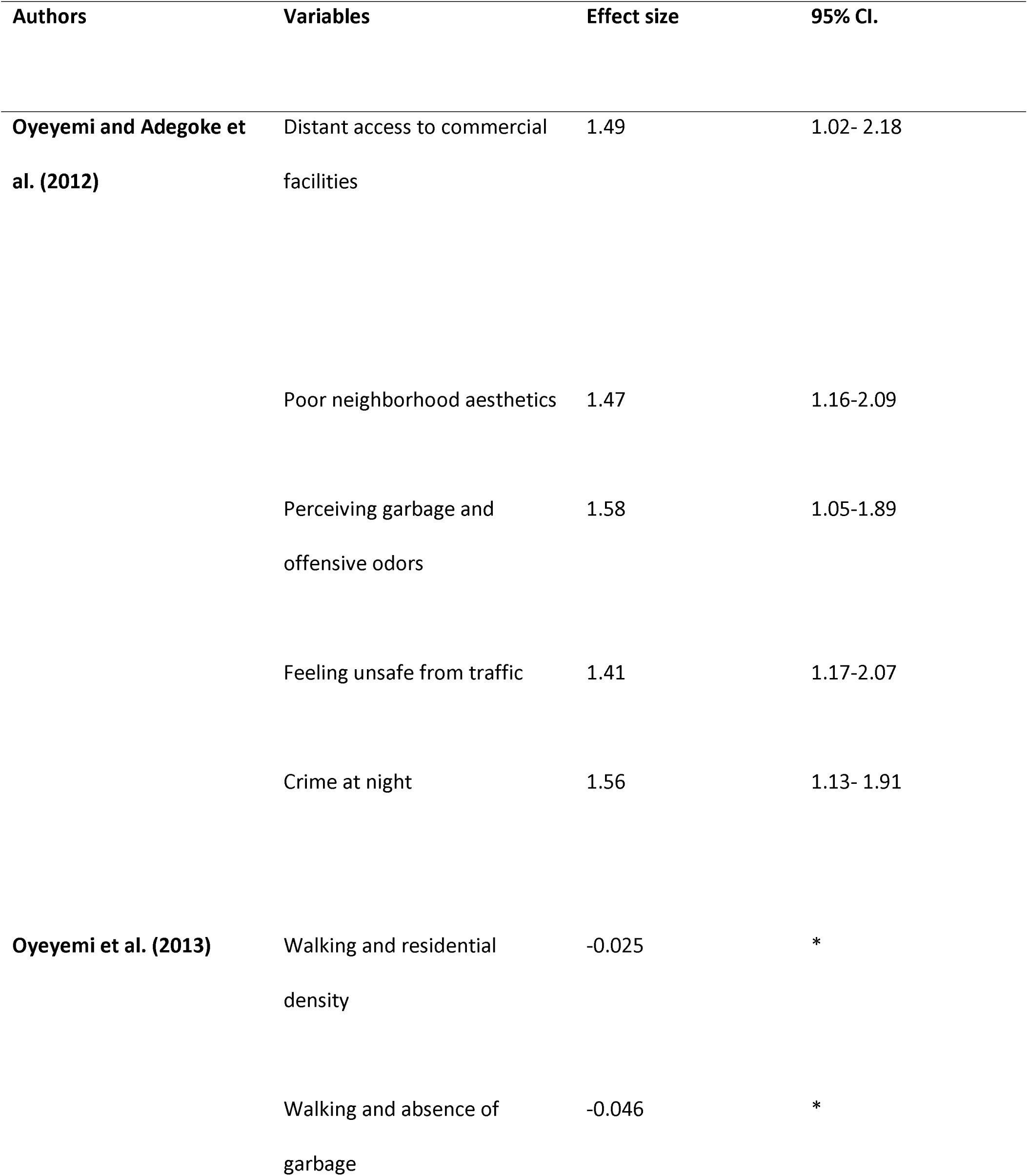
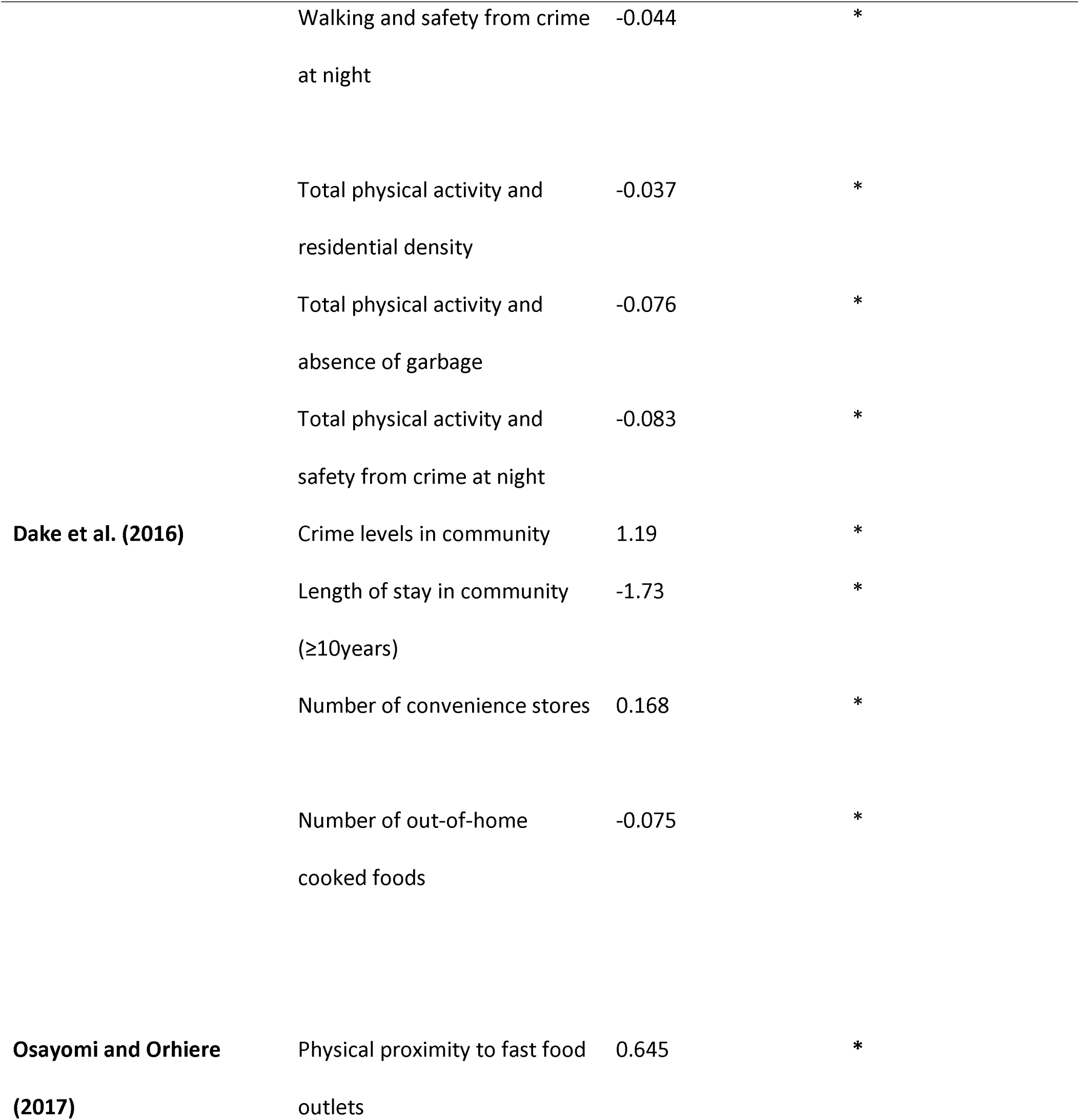
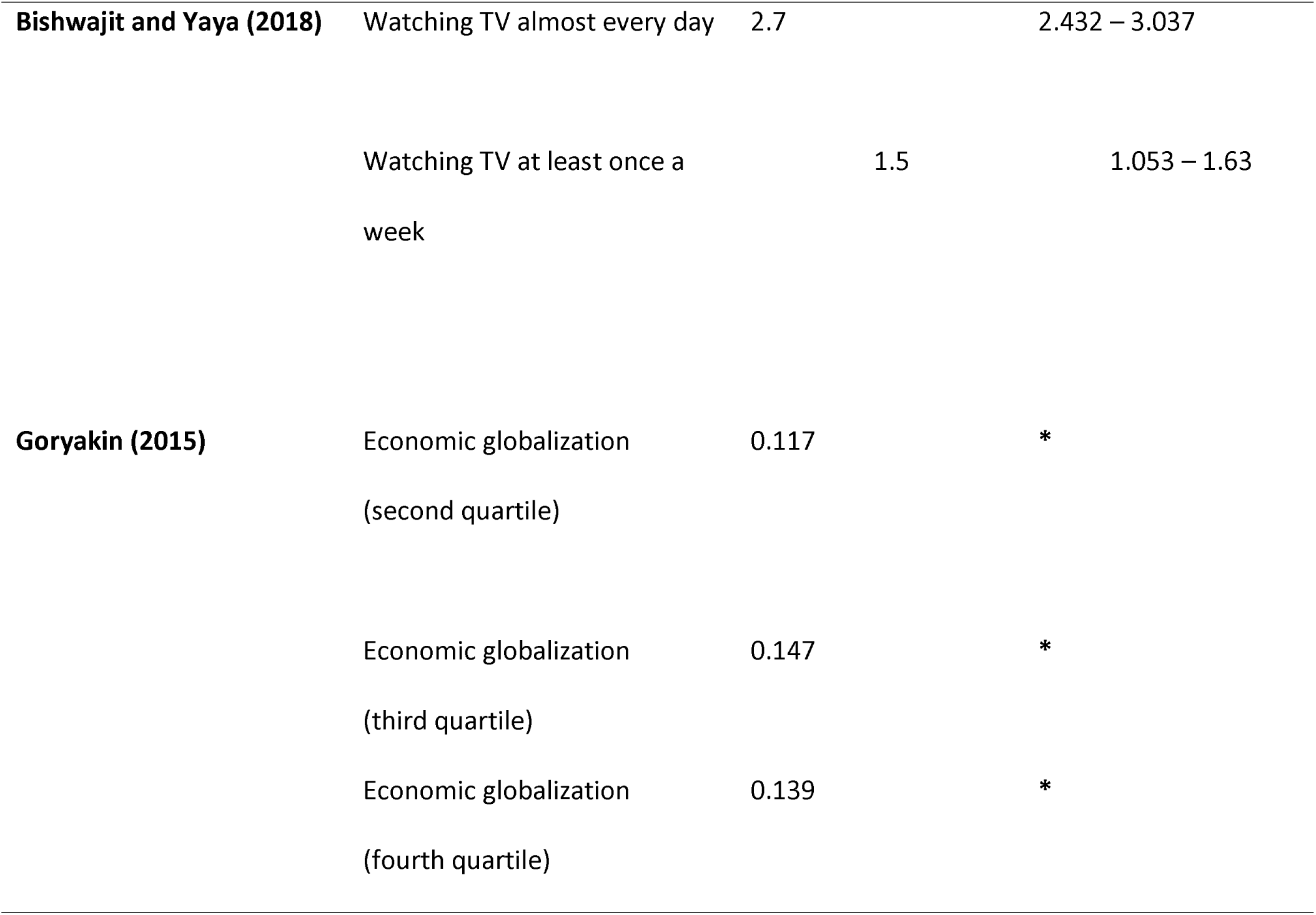
Heterogeneous findings on environmental determinants of obesity

## Discussion

This review aimed to explore the determinants of obesity in West Africa, one of Africa’s subregions currently experiencing a rise in obesity prevalence. Sixty-three studies, mainly from Ghana, Nigeria, Cameroon, Burkina Faso, Cote d’ Ivoire, Senegal, Togo, Benin and Mali were included in this review. The identified determining factors were categorised into demographic, socio-economic, biological, lifestyle and environmental factors. The findings of this review will be discussed based on these categorisations.

In terms of demographic factors, the review identified being a woman and increasing age as common determinants of obesity. One possible explanation for sex differences in obesity determinants is the influence of gonadal steroids on adipose tissue storage and distribution^45^. Evidence from several studies suggests the endocrine mechanism as the cause of the different obesity phenotypic expressions across sex^46–48^. The endocrine pathway predisposes women to have a higher likelihood of having obesity than males in given population. Also, natural processes, like menopause, influence sex variations in obesity prevalence. Women are more inclined to undergo hormonal changes, which can affect glucose regulation, during menopause, and this consequently could predispose them to increased risk of obesity^49, 50^. Regardless of these hormonal explanations, evidence also shows that gender, as a social construct, also predisposes to obesity than men^51, 52^. Women are less likely to engage in physical activities^51, 53^ and more likely to consume foods high in added sugars, such as chocolate and ice-cream than men^54^. However, they are more likely to report healthy eating practices^55^. Additionally, studies indicate that most West African women settle for more sedentary occupations, like petty and table trading; as such, they remain less physically active and subsequently store more body fat^52, 56^. Also, data indicates that in most populations, particularly in LMICs, gender-based food preferences, likely to be influenced by one’s socio-cultural orientation, exists and these could inadvertently result in gender variations in obesity prevalence preferences^55, 57^. Furthermore, studies have shown that most West African communities admire women with large body size^58–60^; thus, most of the women deliberately gain weight to meet these socio-cultural expectations.

The association between increasing age and obesity is also attributed to hormonal/metabolic changes that occur with ageing^61–64^. Ageing is associated with metabolic imbalance, which is a significant contributor to obesity^63^. The increase in perivascular adipose tissue associated with ageing influences the risk of obesity among the aged population^64^. Moreover, ageing is also associated with less physical activities because of musculoskeletal changes, like reduced joint spaces and osteoporosis, that occur with ageing^65^. As most developing countries undergo demographic transition, the increasing burden of conditions, like obesity, is expected^66^. Also, an increase in life expectancy is usually correlated with increase susceptibility with chronic diseases, like diabetes, that is also associated with obesity^63^. Nonetheless, the association between ageing and obesity can go both ways, as obesity can also hasten ageing phenotypic development^67^. These findings on ageing and obesity re-enforce the need for stakeholders to map specific levels of interventions targeted at reducing disease burdens associated with ageing^68^

Education level, employment status and socio-economic status were the identified socio-economic determinants of obesity. Studies^59, 60, 69^ show that large body size is usually tied to affluence and social status in West African countries, like Ghana and Nigeria. Therefore, individuals with high education and employment status may tend to gain weight to keep up with the social status that comes with affluence^70^. Also, wealth usually comes with increased purchasing power, and this inherently increases accessibility to food choices that can be abused^71^. Additionally, most fast-food outlets are in affluent neighbourhoods to attract high-earning clienteles to ensure business viability. This creates obesogenic environments that can be a catalyst for obesity^71^. Furthermore, the centralised systems of most West African countries put residential areas in city peripheries and work environments in city centres^72^. Thus, most people would have to commute long distances to and from work. Consequently, most workers would stay late at work to beat heavy traffic from the centre to the periphery of cities. These could induce stress and result in late eating, factors that have been implicated in the obesity epidemic^72^. The argument on high education and employment status and obesity can also hold for individuals low on the education and employment strata. For such individuals, their food choices are influenced mainly by their affordability^73^. This, however, does not necessarily translate into healthy food choices that could reduce their risk of obesity^74^.

The findings of this review are similar to the review findings of other reviews in both LMICS and HICs^70, 75–79^. However, they were also inconsistent with other review^80^. The review found men to be the common determinant of obesity as opposed to the finding of this review. This difference may be attributed to differences in sample characteristics.

### Policy implications of review findings

The findings of this review indicate the need for specific policies to curtail the obesity epidemic in West Africa and, subsequently, other LMICs. These policies could include implementation of interventions to address gendered eating behaviours and education on socio-cultural weight perceptions and body image from the community-level up to the national level. Also, community-based physical activity interventions, appropriate for all age brackets, could be instituted in schools, churches and other social organisations to address age-related obesity incidence. Additionally, governments, through stakeholder engagements could address obesogenic environments by increasing accessibility and affordability of local foods rich in fibres, minerals, and vitamins, and reduce consumption of foods high in added salts and sugars at all population levels. Furthermore, governments could implement policies, such as decentralisation of government systems and enabling local economies, to reduce rural-urban migration, which is consequential in the obesity menace. Finally, social marketing approaches must be strategised to ensure obesity awareness to elicit favourable changes at individual and environmental levels.

### Strength and limitations of this review

To the best of the researcher’s knowledge, this is the first study to review the determinants of obesity in West Africa. Therefore, the findings could serve as a foundation for other future reviews/studies on obesity determinants in West Africa, in addition to providing a menu of policy options for West African nations and wider LMICs on obesity prevention and control. Also, this review included many studies; hence, it presents with more robust evidence on the determinants of obesity, and this could influence policy shift in the conceptualisation of relevant policies on obesity in West Africa. Even though this review included studies from most West African countries, there was a significant imbalance in the country representation because 65% of the studies were from Nigeria and Ghana. Thus, the evidence on the determinants of obesity in the underrepresented countries is limited. Furthermore, the heterogeneity across the reviewed studies could have influenced the conclusions of this review, and subsequently, the implications as discussed above. Thus, caution must be taken in the interpretation of this review findings to avoid biased inferences.

### Conclusion and Recommendation

The findings of this review indicated that obesity in West Africa is determined by demographic, lifestyle, biological and socio-economic factors such as age, sex, physical activity, and education. These findings present an urgent need for robust engagement with broader stakeholder groups to develop sustainable obesity prevention and control policies to address the obesity epidemic in West Africa. These policies could include education, awareness and implementation of diet and physical activity interventions to stimulate individual and environmental changes at subpopulation and population levels. This review recommends that future studies are conducted in other African subregions, like East Africa, to provide evidence on determinants of obesity to proffer obesity interventions that would have far-reaching benefits to the broader African continent.

## Data Availability

All relevant data are either included in this article or added to the supplementary information.

## Acknowledgements

The authors acknowledge the contributions of the Office of the President (Ghana) and Brunel Global Public Health Academy. The comments herein are those of the authors and not the aforementioned institutions.

